# Integrating genome-wide polygenic risk scores and non-genetic risk to predict colorectal cancer diagnosis: a cohort study in UK Biobank

**DOI:** 10.1101/2021.09.22.21263962

**Authors:** Sarah E.W. Briggs, Philip Law, James E. East, Sarah Wordsworth, Malcolm Dunlop, Richard Houlston, Julia Hippisley-Cox, Ian Tomlinson

## Abstract

**Objectives:** To evaluate the benefit of combining polygenic risk scores (PRS) with the QCancer-10 (colorectal cancer) non-genetic risk prediction model to identify those at highest risk of colorectal cancer (CRC).

**Design:** Population based cohort study. Six different PRS for CRC were developed (using LDpred2 PRS software, clumping and thresholding approaches, and genome-wide significant models). The top-performing genome-wide and GWAS-significant PRS were then combined with QCancer-10 and performance compared to QCancer-10 alone. Case-control (logistic regression) and time-to-event (Cox proportional hazards) analyses were used to evaluate risk model performance in men and women.

**Setting and participants:** UK Biobank Study. A total of 434587 individuals with complete genetic and QCancer-10 predictor data were included in the QCancer-10+PRS modelling cohorts.

**Main outcome measures:** Prediction of colorectal cancer diagnosis by genetic, non-genetic and combined risk models.

**Findings:** PRS derived using the LDpred2 program performed best, with an odds-ratio per standard deviation of 1.58, and top age- and sex-adjusted C-statistic of 0.733 (95% confidence interval 0.710 to 0.753) in logistic regression models in the validation cohort. Integrated QCancer-10+PRS models out-performed QCancer-10 alone. In men, the integrated LDpred2 (QCancer-10+LDP) model produced a C-statistic of 0.730 (0.720 to 0.741) and explained variation of 28.1% (26.3% to 30.0%), compared with 0.693 (0.682 to 0.704) and 21.0% (18.9% to 23.1%) for QCancer-10 alone. Performance improvements in women were similar. In the top 20% of individuals at highest absolute risk, the sensitivity of QCancer-10+LDP models for predicting CRC diagnosis within 5 years was 47.6% in men and 42.5% in women, with respective 3.49-fold and 2.75-fold absolute increases in the top 5% of risk compared to average. Decision curve analysis showed that adding PRS to QCancer-10 improved net-benefit and interventions avoided, across most probability thresholds.

**Conclusions:** Integrating PRS with QCancer-10 significantly improves risk prediction over QCancer-10 alone. Evaluation of risk stratified population screening using this approach is warranted.

**Summary Box:** *What is already known on this topic:*

- Risk stratification based on genetic or environmental risk factors could improve cancer screening outcomes
- No previously published study has examined integrated models combining genome-wide PRS and non-genetic risk factors beyond age
- QCancer-10 (colorectal cancer) is the top-performing non-genetic risk prediction model for CRC

*What this study adds:*

- Adding PRS to the QCancer-10 (colorectal cancer) risk prediction model improves performance and clinical benefit, with greatest gain from the LDpred2 genome-wide PRS, to a level that suggests utility in stratifying CRC screening and prevention

## Introduction

Colorectal cancer (CRC) is the fourth most common cancer in the United Kingdom (UK), with increasing incidence in younger ages and countries with historically lower rates. ^1^ Population screening is effective in reducing CRC incidence and mortality, through detection and removal of pre-malignant adenomas, and earlier detection of cancers. Screening modalities vary internationally. While colonoscopy is the gold-standard, it is expensive, invasive and time consuming. Many countries have adopted a staged process, with initial faecal blood testing, followed by colonoscopy for those who test positive. Risk-stratified approaches to screening direct resources to those at highest risk, have the potential to improve screening detection rates, reduce investigative burden of those at lower risk, and potentially improve cost-effectiveness. ^2^ Improved understanding of cancer risk could also improve informed consent and shared decision making around screening participation.

Both genetic and non-genetic factors contribute to an individual’s risk of CRC, some of the latter being modifiable. The top-performing non-genetic risk model in external validation is QCancer-10 (colorectal cancer), ^3,4^ which has been recommended as a tool to guide shared decision making around CRC screening. ^5^ QCancer-10 is a 15-year CRC prediction model, developed using the QResearch linked primary care database of almost 5 million individuals aged 25-84, registered at QResearch practices across England between 1998 and 2013. ^4^ It is based on age, ethnicity, family history, alcohol and smoking status, a small number of medical conditions, and for men, Townsend deprivation score and body mass index (BMI). As the predictors are derived from electronic health records, it could be embedded at point of care, and linked with screening records to facilitate risk stratification within the bowel screening programme.

Genetic variants known to predispose to CRC are mostly single nucleotide polymorphisms (SNPs) identified as significant in genome-wide association studies (GWAS). Genetic risk can be summarised in a polygenic risk score (PRS). Most existing PRS have used a limited set (typically tens) of risk SNPs that have achieved formal statistical significance in GWAS, with genotypes weighted by predicted effect sizes. ^6^ More recently, “genome-wide PRS” have incorporated many more SNPs than those reaching GWAS-significance, based on the notion that many true risk SNPs remain unidentified. These models have generally produced better performance than “GWAS-significant” models, but evaluation in CRC has been limited. ^7,8^ A further issue is that several previous evaluations of CRC PRS in the UK Biobank Study (UKB) are based on summary statistics derived from a GWAS meta-analyses which included UKB. ^8,9^ This overlap results in overfitting of models, and optimism in performance estimates. ^10^

Integrated models for CRC, which to date have combined GWAS-significant PRS with non-genetic risk factors, generally perform better than non-genetic models or PRS alone. ^6,9^ We hypothesised that integrating PRS with QCancer-10 would provide enhanced risk prediction, and that genome-wide PRS would give greatest benefit. We used UKB to develop and compare PRS using several genome-wide and GWAS-significant approaches, minimising overfitting and optimism by using summary data which did not overlap with the UKB dataset. We validated PRS performance in Geographic and Minority Ethnic Validation Cohorts. We then derived integrated QCancer-10+PRS risk models, using the top-performing genome-wide PRS and the GWAS-significant PRS, which we internally validated and compared with QCancer-10.

## Methods

### Overview

We conducted a development and validation study of PRS and integrated PRS-epidemiological models, to predict risk of CRC in a set of UK individuals of bowel cancer screening age. We followed the PRS-RS and TRIPOD reporting guidelines for PRS and prediction modelling. ^11,12^

We used UKB to derive and validate our risk models, under application number 8508. ^13^ In brief, just over 500,000 participants aged 40 to 69 (5.5% of invitees) were recruited to UKB from the general population across the UK between 2006 and 2010. ^14^ Baseline demographics, medical, lifestyle and physical data, and blood samples, were collected at recruitment. Follow-up through linked hospital, general practice and registry data is ongoing. A detailed description of genetic resources including quality control measures can be found in Bycroft et al. ^13^ and Supplementary Methods. In summary, participants were genotyped on one of two arrays (49950 individuals on the Applied Biosystems UK BiLEVE Axiom Array and the remainder on the Applied Biosystems UK Biobank Axiom Array) which share over 95% content. Following quality control, phasing was carried out using SHAPEIT3 with 1000 Genomes Phase 3 as a reference panel, followed by imputation using IMPUTE4 with the Haplotype Reference Consortium (HRC) dataset as the main reference panel, and secondarily with merged UK10K and 1000 Genomes phase 3 reference panels, and the datasets combined. SNP annotation was based on the GRCh37 assembly of the human genome.

### Outcomes

The primary outcome in all models was CRC diagnosis, identified through self-report at UKB enrolment visit and ICD-9 (153, 154.0, 154.1) and ICD-10 (C18-C20) codes in linked cancer and death registry and hospital data. For PRS development and evaluation in logistic regression models, we included incident and prevalent cases, with the remaining cohort used as controls. For survival analysis using Cox proportional hazards (Cox) models, prevalent cases with a diagnosis prior to cohort entry were excluded. Follow-up began at date of enrolment, and was censored at the earliest of date of incident CRC, loss to follow-up, death, or end of available registry follow-up (31 ^st^ October 2015 for Scottish participants, 13 ^th^ March 2016 for all other participants).

We calculated age-specific and directly standardised CRC incidence rates in UKB overall and for the Integrated Modelling Cohort, and compared these with Office for National Statistics 2013 cancer registry data for England (chosen as the approximate mid-point of available UKB follow-up). ^15^ Age-specific rates were calculated in 5 year age bands between 40 and 80 as the number of first incident CRCs over the number of person years at risk. Age-standardised incidence rates were calculated using the 2013 European Standard Population aged 40-80. Rates are presented per 100000 person years at risk (see Supplementary Methods).

### Polygenic risk scores

We meta-analysed summary data from 14 CRC GWAS cohorts (which did not include UKB) to provide SNP association effect sizes (see Supplementary Methods and Ref.^*16*^). There were 26397 cases and 41481 controls, all of European ancestry based on principal components analysis (PCA). The meta-analysis was performed using the META package (v1.7), ^17^ including SNPs imputed with an imputation quality (INFO) score > 0.8 from each dataset, using the fixed-effects inverse-variance method.

Three broad approaches to PRS development were evaluated (see Supplementary Methods). Firstly, we used a ‘standard’ PRS (hereafter ‘GWAS-sig’), which comprised a manually curated list of 50 sentinel SNPs shown in recent European GWAS-meta-analyses ^16,18^ to be independently and reproducibly associated with CRC risk at *P*<5×10 ^-8^ in our meta-analysis. This PRS was constructed as a log-additive sum of SNP dosages weighted by their betas. Betas were adjusted for winner’s curse using FIQT correction. ^19^

Secondly, genome-wide clumping and thresholding methodologies were evaluated using ‘standard’ (C+T) and ‘stacked’ (SCT) approaches. ^20^

Thirdly, we used LDpred2, ^21^ which takes a Bayesian approach to SNP selection, accounting for linkage disequilibrium between the SNPs. We used three different LDpred2 options – an infinitesimal model (LDpred2-Inf), a non-sparse grid model (LDpred2-grid) and a sparse grid model (LDpred2-grid-sp).

Figure 1 shows the per-person quality control (QC) measures for the genetic data and sample exclusions for each modelling cohort. We used imputed dosage data from UK Biobank, and restricted SNPs to those included in the HapMap3 reference dataset, and with matched SNPs in the base data. Following QC, 1104409 SNPs were available for PRS development (Supplementary Methods).

**Figure 1.**
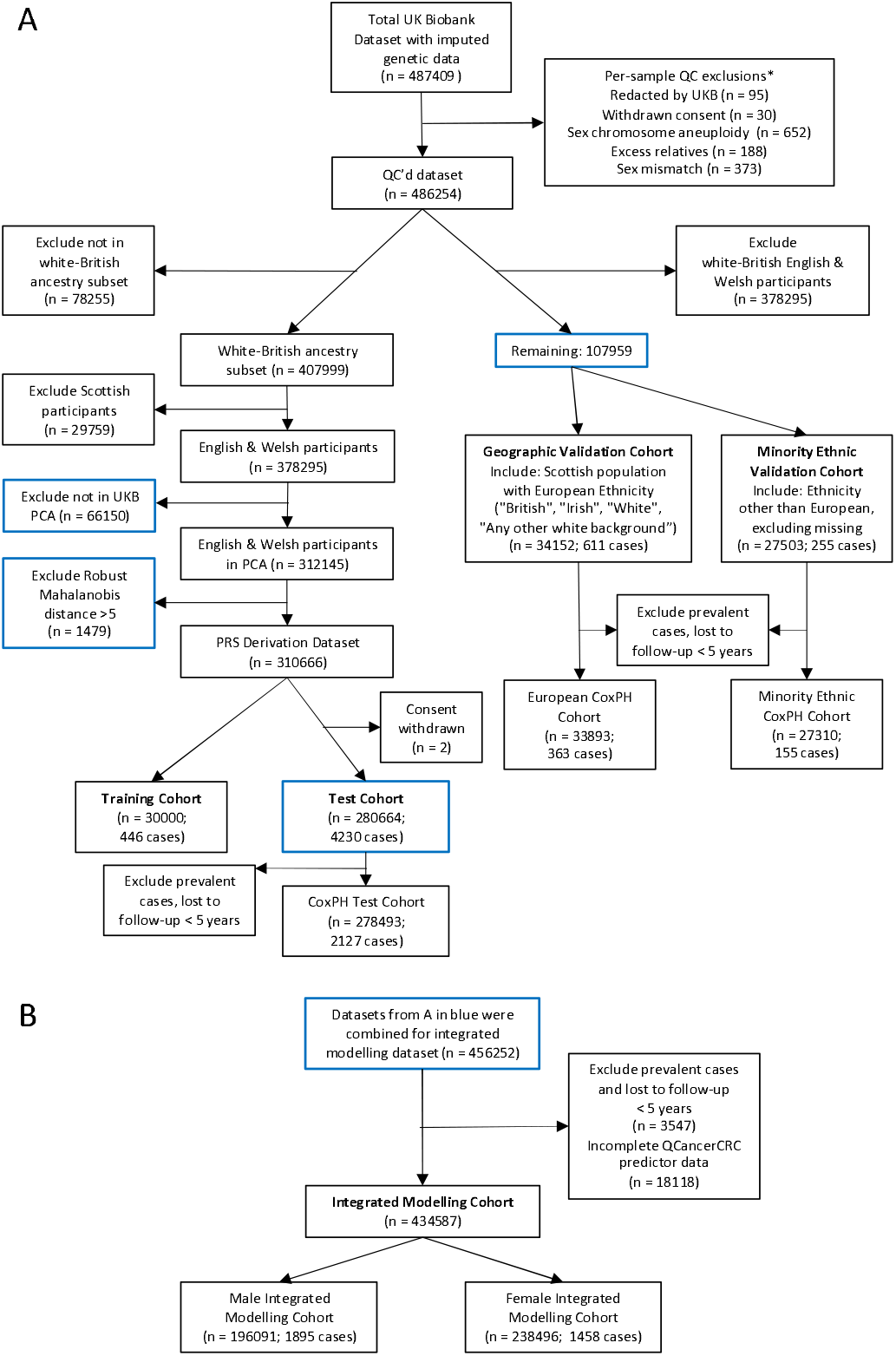
UK Biobank participant flow diagram. Panel A shows quality control and derivation of PRS modelling cohorts. Panel B shows participant selection for the Integrated Modelling Cohorts. *More than one exclusion may apply per person.

Optimal PRS tuning parameters for genome-wide approaches were selected in the Training Cohort (n=30000, 446 cases; Supplementary Methods, Table S4). For each optimal PRS, we assessed association with CRC risk in logistic regression and Cox risk models in the Test cohort (n=280664; 4230 cases), adjusting for age, sex, genotyping array and the first four principal components (PCs) from UKB. We tested for interactions between age and PRS. Training and Test Cohorts included participants of white-British ancestry (identified through self-reported ethnicity and genetic information) ^13^ from England and Wales (Figure 1). Case prevalence was 1.5% in both cohorts. We compared performance to a reference model containing age, sex, genotyping array and four PCs, without the PRS. We also evaluated performance without age and sex in the model.

We reported the distribution of standardised PRS and adjusted odd ratios and hazard ratios per-standard deviation (Supplementary Methods). We used the C-statistic (Harrell’s C-index for Cox models) and Somers’ D_xy_ statistic to assess discrimination, in addition to Royston’s D statistic and separation of Kaplan-Meier curves across four risk groups (cut at 16th, 50th and 84th centiles, approximating to the mean and 1 standard deviation) ^22^ for Cox models. Nagelkerke’s *R*^2^ was used in logistic regression models and Royston and Sauerbrei’s 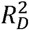 in Cox models to assess variance explained, and *R*^2^ attributable to the PRS was calculated by *R*^2^ (full model) *R*^2^-(reference model). These measures were evaluated over the follow-up time of the cohort for Cox models. Scaled Brier scores (derived from the Brier score scaled to the maximum possible score for a given dataset, where a higher % score indicates better performance), ^23^ were used to assess overall model performance, calculated at 8 years of follow-up for Cox models. Each model was internally validated. Confidence intervals and internal validation used 500 bootstrap samples.

Prior to external validation, models were adjusted for optimism. The optimism-adjusted calibration slope was used as a global shrinkage factor to adjust the regression coefficients, and the intercept or baseline survival function was re-estimated (by refitting the model with the adjusted linear predictor as an offset). ^24^ Adjusted PRS models were then applied to a Geographic Validation Cohort, comprising Scottish participants with European ancestry, and a Minority Ethnic Validation Cohort (from any region). The null hypothesis of no difference in performance statistics between models was tested using paired t-tests with Bonferroni correction for multiple comparisons. In addition to the performance metrics described above, calibration was assessed through the calibration slope and visual assessment of calibration plots, with calibration-in-the-large for logistic regression models. Pre-specified subgroup analyses were performed in the Geographic Validation Cohort by sex, in those with a first degree family history of CRC, and by age (Supplementary Methods). We evaluated potential improvements in calibration in validation datasets obtained through recalibration-in-the-large (in which the intercept or baseline survival function is re-estimated in the new dataset).

### Development of QCancer-10+PRS combined models

Coding of QCancer-10 predictors in UK Biobank were matched as closely as possible to the original model. ^4^ Ethnicity, previous medical history, alcohol and smoking status, and family history were all obtained from self-reported data in baseline touch-screen responses and verbal interviews at UKB assessment centres. Mapping of QCancer-10 predictors to UK Biobank data and coding of predictors is described in Supplementary Methods and Tables S1 and S2.

The Integrated Modelling Cohort used for QCancer-10 validation and integrated model development comprised all individuals with imputed genetic data passing QC, excluding the 30000 individuals used for PRS hyper-parameter selection (Supplementary Methods), and with complete QCancer-10 predictor data (Figure 1). Since missingness was <5% for all predictors (Table S3), we used complete case analysis. Sample size adequacy for integrated model development was calculated following Riley *et al*. ^25^ (Supplementary Methods).

We validated QCancer-10 performance in UKB, and recalibrated the model for the UKB dataset through recalibration-in-the-large. Full QCancer-10 model specification is available at https://www.qcancer.org/15yr/colorectal/. We then developed integrated models including the risk score from QCancer-10 plus either the top-performing genome-wide PRS (based on the maximum C-statistic and *R*^2^ in external validation) or the GWAS-significant PRS, using Cox models, developed in men and women separately. Inspection of Schoenfeld residuals showed that the proportional hazard assumption held. We evaluated the use of multiple fractional polynomials to model the predictors, ultimately using a fractional polynomial term to model the genome-wide PRS in the model for females. We assessed for possible interactions between the predictors by visual inspection of plots of marginal effects of the QCancer-10 risk score across PRS values, and examining the prognostic strength and significance of interaction terms based on Wald *χ*^2^statistics.

We used the same metrics to assess the original QCancer-10 model and QCancer-10+PRS model performance as described for Cox PRS models, with paired t-tests to compare models as above. Confidence intervals and internal validation used 500 bootstrap samples. Pre-specified subgroup analyses for QCancer-10 and QCancer-10+PRS included those with a first degree family history of CRC, individuals from minority ethnic backgrounds, and calibration by age.

Model sensitivities were evaluated by calculating the proportion of cases identified at centile thresholds for absolute risk and relative risk. Relative risks were calculated relative to an individual of the same age and sex, mean PRS (by sex), mean PCs, BMI of 25, white ethnicity, mean Townsend score, and no other CRC risk factors. We used decision curve analyses to compare the net benefit and interventions avoided using QCancer-10 and QCancer-10+PRS models. For decision curve and subgroup analyses, QCancer-10+PRS models were first adjusted for optimism, and recalibrated QCancer-10 models were used.

Statistical analysis was performed using R/3.6.2. ^26^

### Patient and Public Involvement

Patients and public were not involved in the design, conduct, reporting or dissemination plans of this study.

## Results

Demographics for the UKB-derived Integrated Modelling Cohort are shown in Table 1. Table S3 gives these values, including numbers not reported, for the whole UKB cohort; characteristics of each PRS cohort are shown in Table S4. Age-standardised CRC incidence from linked cancer-registry data in the whole UKB cohort was 108.3 and 73.9 cases per 100000 person years at risk for men and women respectively, compared to 127.8 and 80.7 respectively in ONS data. ^15^ Incidence in the Integrated Modelling Cohort, with cases identified through all linked data, was 118.0 CRCs per 100,000 years follow-up in men and 79.3 in women. Age-specific incidence rates in UKB (Figure S3) closely followed those from ONS until the age of 70, after which UKB rates were lower.

**Table 1.**
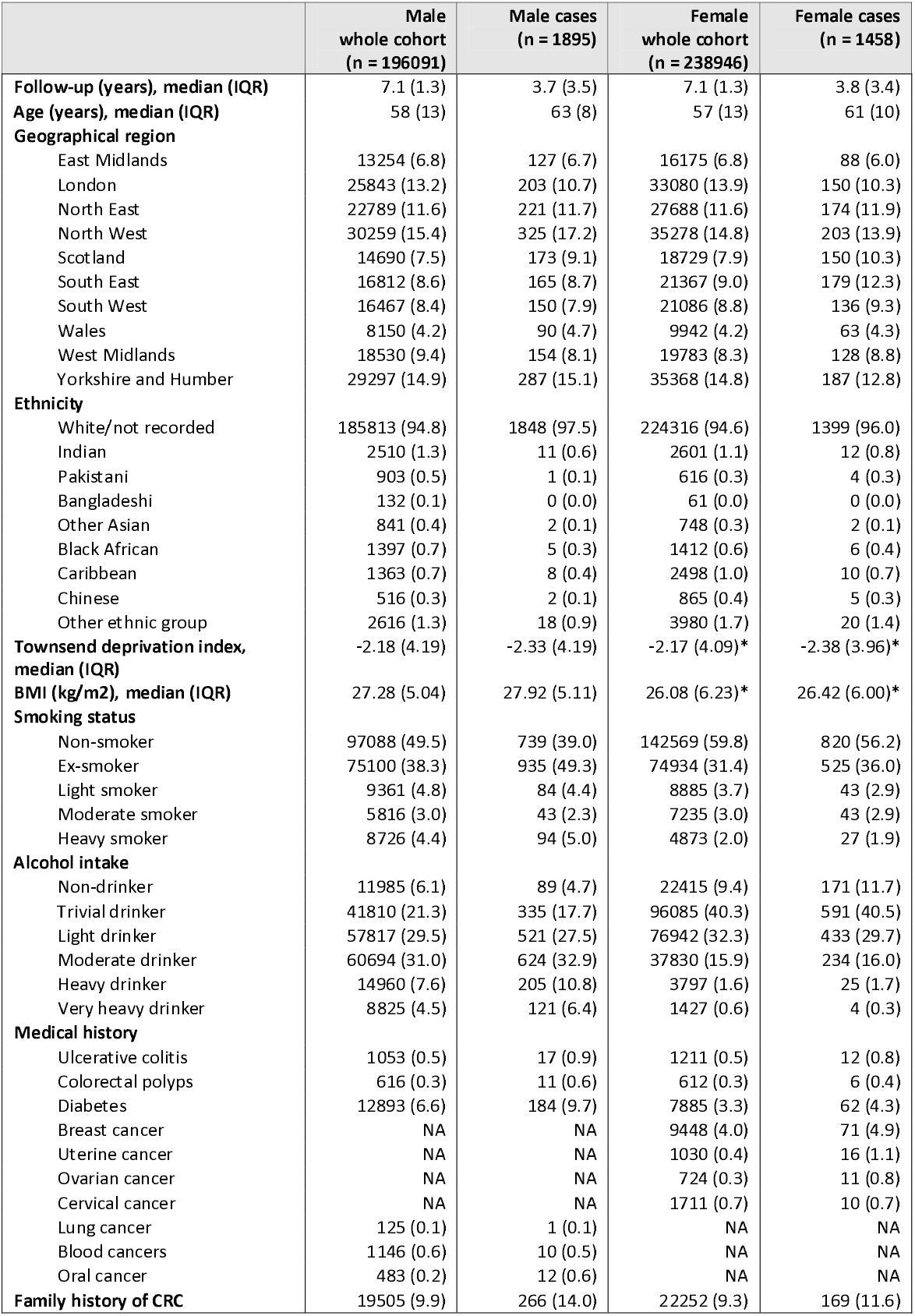
Demographic data and medical conditions included in QCancer-10 models, in male and female Integrated Modelling Cohorts, and in cases. Values are numbers (%) unless otherwise indicated. CRC – colorectal cancer, IQR – interquartile range, NA – not applicable. *not included in model for females but provided for information.

|  | Male<br>whole cohort<br>(n = 196091) | Male cases<br>(n = 1895) | Female<br>whole cohort<br>(n = 238946) | Female cases<br>(n = 1458) |
| --- | --- | --- | --- | --- |
| **Follow-up (years), median (IQR)** | 7.1 (1.3) | 3.7 (3.5) | 7.1 (1.3) | 3.8 (3.4) |
| **Age (years), median (IQR)** | 58 (13) | 63 (8) | 57 (13) | 61 (10) |
| **Geographical region** |  |  |  |  |
| East Midlands | 13254 (6.8) | 127 (6.7) | 16175 (6.8) | 88 (6.0) |
| London | 25843 (13.2) | 203 (10.7) | 33080 (13.9) | 150 (10.3) |
| North East | 22789 (11.6) | 221 (11.7) | 27688 (11.6) | 174 (11.9) |
| North West | 30259 (15.4) | 325 (17.2) | 35278 (14.8) | 203 (13.9) |
| Scotland | 14690 (7.5) | 173 (9.1) | 18729 (7.9) | 150 (10.3) |
| South East | 16812 (8.6) | 165 (8.7) | 21367 (9.0) | 179 (12.3) |
| South West | 16467 (8.4) | 150 (7.9) | 21086 (8.8) | 136 (9.3) |
| Wales | 8150 (4.2) | 90 (4.7) | 9942 (4.2) | 63 (4.3) |
| West Midlands | 18530 (9.4) | 154 (8.1) | 19783 (8.3) | 128 (8.8) |
| Yorkshire and Humber | 29297 (14.9) | 287 (15.1) | 35368 (14.8) | 187 (12.8) |
| **Ethnicity** |  |  |  |  |
| White/not recorded | 185813 (94.8) | 1848 (97.5) | 224316 (94.6) | 1399 (96.0) |
| Indian | 2510 (1.3) | 11 (0.6) | 2601 (1.1) | 12 (0.8) |
| Pakistani | 903 (0.5) | 1 (0.1) | 616 (0.3) | 4 (0.3) |
| Bangladeshi | 132 (0.1) | 0 (0.0) | 61 (0.0) | 0 (0.0) |
| Other Asian | 841 (0.4) | 2 (0.1) | 748 (0.3) | 2 (0.1) |
| Black African | 1397 (0.7) | 5 (0.3) | 1412 (0.6) | 6 (0.4) |
| Caribbean | 1363 (0.7) | 8 (0.4) | 2498 (1.0) | 10 (0.7) |
| Chinese | 516 (0.3) | 2 (0.1) | 865 (0.4) | 5 (0.3) |
| Other ethnic group | 2616 (1.3) | 18 (0.9) | 3980 (1.7) | 20 (1.4) |
| **Townsend deprivation index, median (IQR)** | -2.18 (4.19) | -2.33 (4.19) | -2.17 (4.09)* | -2.38 (3.96)* |
| **BMI (kg/m<sup>2</sup>), median (IQR)** | 27.28 (5.04) | 27.92 (5.11) | 26.08 (6.23)* | 26.42 (6.00)* |
| **Smoking status** |  |  |  |  |
| Non-smoker | 97088 (49.5) | 739 (39.0) | 142569 (59.8) | 820 (56.2) |
| Ex-smoker | 75100 (38.3) | 935 (49.3) | 74934 (31.4) | 525 (36.0) |
| Light smoker | 9361 (4.8) | 84 (4.4) | 8885 (3.7) | 43 (2.9) |
| Moderate smoker | 5816 (3.0) | 43 (2.3) | 7235 (3.0) | 43 (2.9) |
| Heavy smoker | 8726 (4.4) | 94 (5.0) | 4873 (2.0) | 27 (1.9) |
| **Alcohol intake** |  |  |  |  |
| Non-drinker | 11985 (6.1) | 89 (4.7) | 22415 (9.4) | 171 (11.7) |
| Trivial drinker | 41810 (21.3) | 335 (17.7) | 96085 (40.3) | 591 (40.5) |
| Light drinker | 57817 (29.5) | 521 (27.5) | 76942 (32.3) | 433 (29.7) |
| Moderate drinker | 60694 (31.0) | 624 (32.9) | 37830 (15.9) | 234 (16.0) |
| Heavy drinker | 14960 (7.6) | 205 (10.8) | 3797 (1.6) | 25 (1.7) |
| Very heavy drinker | 8825 (4.5) | 121 (6.4) | 1427 (0.6) | 4 (0.3) |
| **Medical history** |  |  |  |  |
| Ulcerative colitis | 1053 (0.5) | 17 (0.9) | 1211 (0.5) | 12 (0.8) |
| Colorectal polyps | 616 (0.3) | 11 (0.6) | 612 (0.3) | 6 (0.4) |
| Diabetes | 12893 (6.6) | 184 (9.7) | 7885 (3.3) | 62 (4.3) |
| Breast cancer | NA | NA | 9448 (4.0) | 71 (4.9) |
| Uterine cancer | NA | NA | 1030 (0.4) | 16 (1.1) |
| Ovarian cancer | NA | NA | 724 (0.3) | 11 (0.8) |
| Cervical cancer | NA | NA | 1711 (0.7) | 10 (0.7) |
| Lung cancer | 125 (0.1) | 1 (0.1) | NA | NA |
| Blood cancers | 1146 (0.6) | 10 (0.5) | NA | NA |
| Oral cancer | 483 (0.2) | 12 (0.6) | NA | NA |
| **Family history of CRC** | 19505 (9.9) | 266 (14.0) | 22252 (9.3) | 169 (11.6) |

### Polygenic risk score models

Of the 6 PRS models assessed (Figure S4), LDpred2-grid had the highest ORs per SD of PRS (1.584, 95% confidence interval 1.536 to 1.633; Table 2) and performed best in the test cohort with C-statistic 0.717 (0.711 to 0.725) and R2 6.3% (5.9 to 6.8%) (Table 2). A weak interaction between age and PRS was noted with a reduced effect size of PRS with increasing age (Table S5, Figure S5), but was not included in the models. Genome-wide models performed better than the GWAS-sig model, and all PRS showed improved performance over the reference model of age, sex, genotyping array and four PCs (Table 2). Performance without adjustment for age and sex is shown in Table S6. Internal validation showed low bias in all measures (Table 2).

**Table 2.** Apparent, internally and externally validated polygenic risk score (PRS) performance in logistic regression models (adjusting for age, sex, genotyping array and first 4 principal components). Values are performance indices plus 95% confidence intervals. Internal validation used 500 bootstrap samples. LDpred2-inf – LDpred2 infinitesimal model; LDpred2-grid – LDpred2 grid model; LDpred2-grid-sp – LDpred2 sparse grid model; SCT – stacked clumping and thresholding; C+T – clumping and thresholding; GWAS-sig – GWAS significant; PRS OR per SD – odds ratio per standard deviation of polygenic risk score in the age- and sex-adjusted model; C – C statistic; Dxy – Somers’ Dxy rank correlation; R2 – Nagelkerke’s *R*^2^(explained variation); Slope – Calibration Slope; CITL – calibration-in-the-large. * *R*^2^ for all models in the Minority Ethnic Validation Cohort <0 (indicating poorer performance than a model with no explanatory variables). Pairwise comparisons of performance metrics in validation cohorts were all significantly different P<0.001 except comparisons marked ^#^P=0.002, ^##^P=0.005, ^P=0.01, *P=1.

|  | LDpred2-inf | LDpred2-grid | LDpred2-grid-sp | SCT | C+T | GWAS-sig | Reference |
| --- | --- | --- | --- | --- | --- | --- | --- |
| Number of SNPs | 1,104,409 | 1,104,409 | 616,956 | 194,756 | 306,912 | 50 | NA |
| **Apparent performance** |  |  |  |  |  |  |  |
| PRS OR per SD | 1.435 (1.391 - 1.480) | 1.584 (1.536 - 1.633) | 1.571 (1.524 - 1.620) | 1.417 (1.375 - 1.461) | 1.425 (1.382 - 1.470) | 1.390 (1.348 - 1.433) | NA |
| C | 0.704 (0.697 - 0.712) | 0.717 (0.711 - 0.725) | 0.716 (0.710 - 0.723) | 0.702 (0.695 - 0.711) | 0.704 (0.697 - 0.711) | 0.700 (0.693 - 0.707) | 0.680 (0.672 - 0.687) |
| Dxy | 0.407 (0.394 - 0.423) | 0.435 (0.422 - 0.451) | 0.432 (0.419 - 0.446) | 0.404 (0.389 - 0.422) | 0.407 (0.394 - 0.423) | 0.400 (0.386 - 0.414) | 0.359 (0.344 - 0.374) |
| R2 (%) | 5.5 (5.1 - 5.9) | 6.3 (5.9 - 6.8) | 6.2 (5.8 - 6.7) | 5.4 (5.0 - 5.9) | 5.4 (5.1 - 5.9) | 5.3 (4.9 - 5.7) | 4.2 (3.8 - 4.6) |
| R2 PRS (%) | 1.3 (1.1 - 1.5) | 2.1 (1.9 - 2.4) | 2.0 (1.8 - 2.3) | 1.2 (1.0 - 1.4) | 1.2 (1.0 - 1.5) | 1.1 (0.9 - 1.3) | NA |
| Scaled Brier (%) | 0.87 | 1.05 | 1.03 | 0.86 | 0.85 | 0.83 | 0.60 |
| **Internal validation** |  |  |  |  |  |  |  |
| C | 0.703 | 0.717 | 0.716 | 0.701 | 0.703 | 0.700 | 0.679 |
| Dxy | 0.406 | 0.434 | 0.432 | 0.403 | 0.406 | 0.400 | 0.358 |
| R2 (%) | 5.4 | 6.3 | 6.2 | 5.4 | 5.4 | 5.3 | 4.2 |
| Slope | 0.996 | 0.997 | 0.998 | 0.996 | 0.995 | 0.999 | 0.996 |
| Scaled Brier (%) | 0.85 | 0.94 | 1.06 | 0.84 | 0.76 | 0.85 | 0.58 |
| **Geographic Validation** |  |  |  |  |  |  |  |
| C | 0.726 (0.704 - 0.748) | 0.732 (0.710 - 0.752) | 0.733 (0.710 - 0.753) | 0.718 (0.696 - 0.739)<sup>^</sup> | 0.719 (0.696 - 0.740)<sup>^</sup> | 0.703 (0.679 - 0.724) | 0.677 (0.654 - 0.699) |
| Dxy | 0.452 (0.408 - 0.496) | 0.464 (0.420 - 0.504) | 0.466 (0.421 - 0.507) | 0.436 (0.392 - 0.477)<sup>^</sup> | 0.438 (0.392 - 0.480)<sup>^</sup> | 0.405 (0.358 - 0.447) | 0.353 (0.308 - 0.397) |
| R2 (%) | 7.0 (5.7 - 8.4) | 7.6 (6.1 - 8.9) | 7.6 (6.1 - 8.9) | 6.4 (5.0 - 7.7) | 6.6 (5.2 - 7.9) | 5.4 (4.0 - 6.7) | 3.8 (2.6 - 5.0) |
| Slope | 1.137 (1.010 - 1.268) | 1.091 (0.967 - 1.199) | 1.104 (0.980 - 1.213) | 1.076 (0.946 - 1.200) | 1.098 (0.958 - 1.222) | 0.994 (0.861 - 1.113) | 0.936 (0.795 - 1.075) |
| CITL | 0.206 (0.120 - 0.272) | 0.198 (0.113 - 0.262) | 0.199 (0.115 - 0.264) | 0.194 (0.107 - 0.260) | 0.195 (0.110 - 0.261) | 0.191 (0.104 - 0.258)<sup>*</sup> | 0.191 (0.105 - 0.257)<sup>*</sup> |
| Scaled Brier (%) | 1.48 | 1.64 | 1.66 | 1.27 | 1.38 | 1.08 | 0.67 |
| **Minority Ethnic Validation\*** |  |  |  |  |  |  |  |
| C | 0.588 (0.545 - 0.627) | 0.602 (0.558 - 0.640)<sup>##</sup> | 0.601 (0.559 - 0.640)<sup>##</sup> | 0.589 (0.546 - 0.626) | 0.597 (0.554 - 0.636) | 0.587 (0.543 - 0.624) | 0.585 (0.542 - 0.623) |
| Dxy | 0.176 (0.090 - 0.254) | 0.203 (0.116 - 0.279)<sup>##</sup> | 0.203 (0.118 - 0.281)<sup>##</sup> | 0.179 (0.093 - 0.253) | 0.195 (0.108 - 0.271) | 0.174 (0.086 - 0.247) | 0.171 (0.084 - 0.245) |
| Slope | 0.175 (0.096 - 0.258) | 0.204 (0.122 - 0.288) | 0.208 (0.126 - 0.294) | 0.161 (0.088 - 0.240) | 0.195 (0.110 - 0.281) | 0.143 (0.071 - 0.213)<sup>#</sup> | 0.144 (0.069 - 0.217)<sup>#</sup> |
| CITL | 1.299 (1.155 - 1.417) | 1.336 (1.194 - 1.456) | 1.325 (1.183 - 1.446) | 1.360 (1.217 - 1.479) | 1.310 (1.167 - 1.429) | 1.392 (1.251 - 1.511) | 1.343 (1.200 - 1.459) |
| Scaled Brier (%) | -0.02 | 0.04 | 0.03 | -0.06 | -0.06 | -0.07 | -0.15 |

In the Geographic Validation Cohort, discrimination and variation explained improved compared to the Test Cohort for all models. LDpred2-derived models performed best, and all genome-wide models showed improved performance over the GWAS-sig model (Table 2). All models under-predicted risk (CITL >0, Table 2) particularly in the highest PRS groups (Figure S6), and genome-wide models were slightly over-fitted (calibration slope >1, i.e. insufficient variation at the extremes of prediction, Table 2, Figure S6).

In subgroup analyses of logistic regression models (Table S7, Figure S7), discrimination and explained variation were better in males; models were better fitted in females but under-predicted risk to a greater extent, particularly in higher risk groups. Discrimination and variation explained were poorer in individuals with a first-degree family history of CRC, with models systematically underpredicting risk across PRS risk groups. All models tended to under-predict risk across age groups (Figure S8).

Performance was poor in the Minority Ethnic Validation Cohort (Table 2). Models systematically under-predicted risk and were highly over-fitted (i.e. predictions were too extreme, Table 2), with modest improvement following recalibration (Figure S6).

In general, PRS performance in Cox models supported the logistic regression analysis (Tables S8-9, Figures S9-14).

### QCancer-10 non-genetic model

QCancer-10 models risk in males and females separately. Comparative demographics of the original QCancer-10 derivation cohort and the Integrated Modelling Cohort are shown in Table S10. Notably the Integrated Modelling Cohort is older, less ethnically diverse, has a lower Townsend score, fewer smokers, and higher reported family history of CRC, than the QCancer-10 cohort. Model performance was in line with previously published studies (Table 3). ^3^ As expected, the model for females performed less well than the model for males. ^3^ Both models tended to over-predict risk, which was corrected through recalibration, though in women the model continued to over-predict in the top risk decile (Figure S15). In subgroup analysis, models were well calibrated across age groups; they underpredicted risk in individuals from minority ethnic backgrounds; and the model for females tended to over-predict risk in those with a first-degree family history of CRC, particularly in higher risk groups (Table S11, Figures S16-S17).

**Table 3.** Apparent and internally validated performance of QCancer-10+LDP and QCancer-10+GWS models, compared with external validation of QCancer-10 in the same participants. Values are performance indices plus 95% confidence intervals. QCancer-10/PRS HR per SD – adjusted hazard ratio of QCancer-10 score or PRS in model per standard deviation of the PRS; C – Harrell’s C statistic; Dxy – Somers’ Dxy rank correlation; D – Royston’s D statistic; R2D – Royston and Sauerbrei’s 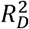 (explained variation); Slope – Calibration Slope. Pairwise comparisons of performance metrics were all significantly different P<0.001. *modelled using multiple fractional polynomial and therefore not presented.

|  | QCancer-10+LDP |  | QCancer-10+GWS |  | QCancer-10 |
| --- | --- | --- | --- | --- | --- |
|  | Apparent | Internal Validation | Apparent | Internal Validation |  |
| Males |  |  |  |  |  |
| QCancer-10 HR per SD | 2.309 (2.164 - 2.464) | NA | 2.334 (2.187 - 2.490) | NA | NA |
| PRS HR per SD | 1.592 (1.520 - 1.668) | NA | 1.453 (1.388 - 1.521) | NA | NA |
| C | 0.730 (0.720 - 0.741) | 0.730 | 0.717 (0.707 - 0.727) | 0.716 | 0.693 (0.682 - 0.704) |
| Dxy | 0.460 (0.440 - 0.481) | 0.460 | 0.433 (0.414 - 0.455) | 0.433 | 0.847 (0.841 - 0.852) |
| D | 1.282 (1.224 - 1.341) | 1.280 | 1.209 (1.156 - 1.267) | 1.207 | 1.058 (0.987 - 1.121) |
| R2D (%) | 28.2 (26.3 - 30.0) | 28.1 | 25.9 (24.2 - 27.7) | 25.8 | 21.1 (18.9 - 23.1) |
| Scaled Brier (%) | 0.81 | 0.80 | 0.81 | 0.80 | 0.59 |
| Slope | NA | 0.998 | NA | 0.998 | 0.995 (0.914 - 1.063) |
| Females |  |  |  |  |  |
| QCancer-10 HR per SD | * | NA | 1.764 (1.655 - 1.881) | NA | NA |
| PRS HR per SD | * | NA | 1.359 (1.290 - 1.431) | NA | NA |
| C | 0.686 (0.672 - 0.701) | 0.685 | 0.668 (0.655 - 0.683) | 0.668 | 0.645 (0.631 - 0.659) |
| Dxy | 0.372 (0.345 - 0.402) | 0.37 | 0.337 (0.310 - 0.366) | 0.335 | 0.822 (0.816 - 0.830) |
| D | 1.056 (0.979 - 1.136) | 1.055 | 0.925 (0.850 - 1.000) | 0.924 | 0.769 (0.695 - 0.847) |
| R2D (%) | 21.0 (18.6 - 23.5) | 21 | 17.0 (14.7 - 19.3) | 16.9 | 12.4 (10.3 - 14.6) |
| Scaled Brier (%) | 0.34 | 0.34 | 0.29 | 0.28 | 0.20 |
| Slope | NA | 0.996 | NA | 0.996 | 0.805 (0.724 - 0.899) |

### QCancer-10+PRS models

Given the similarities in performance of LDpred2-grid and LDpred2-grid-sp models, we selected LDpred2-grid-sp as the top-performing genome-wide PRS for integrated modelling with QCancer-10, favouring sparsity (i.e. a PRS containing fewer SNPs; see Supplementary Results for full model specifications and baseline hazards). We found evidence of an interaction between QCancer-10 and the GWAS-significant PRS in men (P interaction = 0.004), with reduced effect of QCancer-10 score at higher PRS, but did not ultimately include this in the model given the relative weakness of the interaction (Table S12, Figure S18).

Cox models combining the QCancer-10 risk score with LDpred2-grid-sp (QCancer-10+LDP), and the GWAS-sig PRS (QCancer-10+GWS) both out-performed QCancer-10 (Table 3). Figure 2 shows Kaplan-Meier curves across 4 risk groups in integrated QCancer-10+PRS models compared to QCancer-10 alone, demonstrating improved separation between risk groups with the addition of PRS. Internal validation of the QCancer-10+PRS models showed very little optimism in performance estimates.

**Figure 2.**
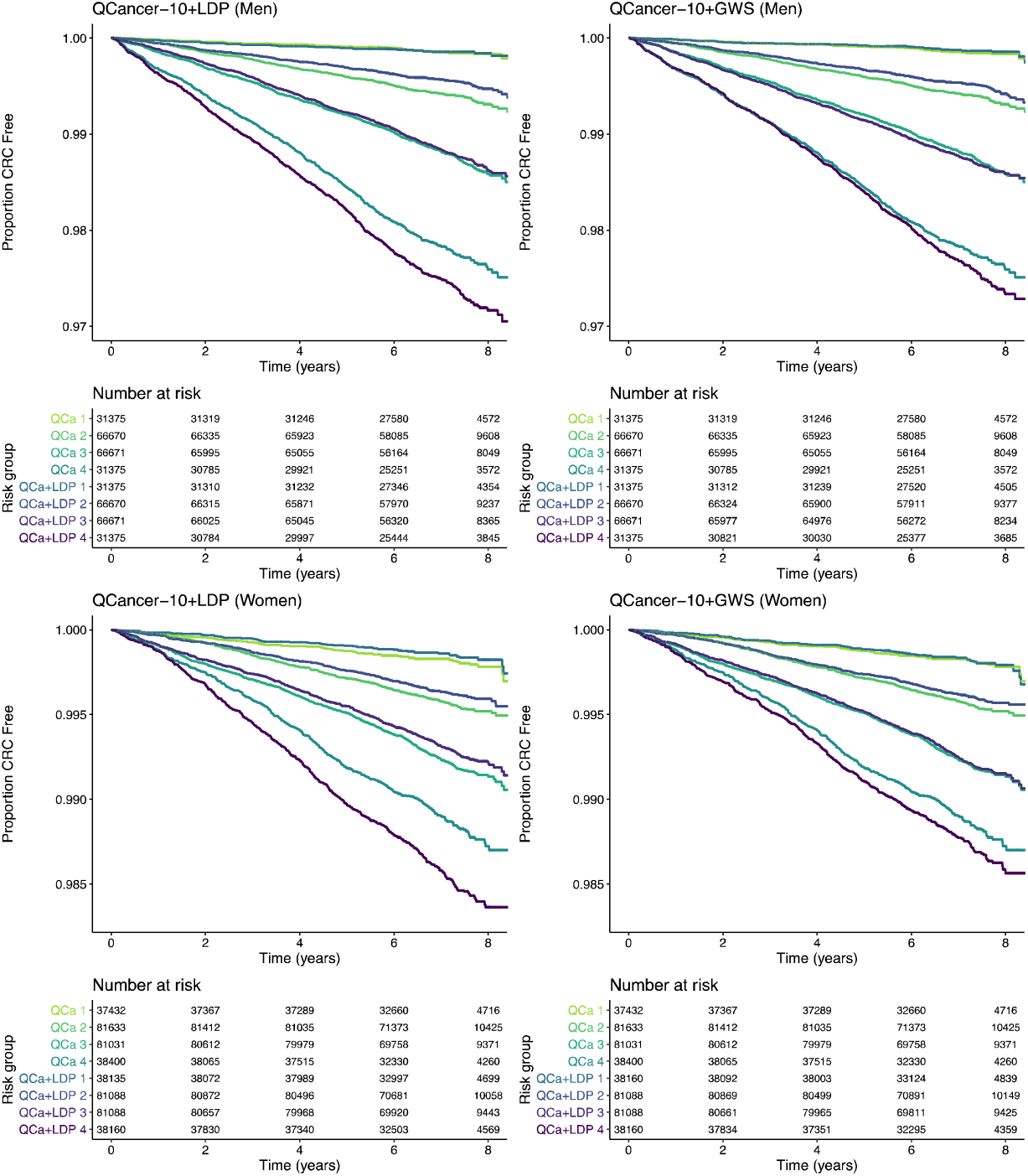
Kaplan-Meier curves across four risk groups (group 4 being highest risk) for QCancer-10+LDP and QCancer-10+GWS models compared to QCancer-10 in men and women. QCa – QCancer-10 model; QCa+LDP – Qcancer-10+LDP model; QCa+GWS – QCancer-10+GWS model.

Models predicting risk in men had better discrimination, and explained more of the variation in risk than models for women (Table 3). Calibration by age was good (Figure S16), with slight under-prediction of risk in the top age group in women. As with QCancer-10, in those with a first degree family history of CRC, female QCancer-10+PRS models tended to over-predict risk, particularly in higher risk groups; male QCancer-10+PRS models were well calibrated (Table S11, Figure S17). In minority ethnicities, QCancer-10+PRS models underpredicted risk (Table S11) to a greater extent than QCancer-10, subject to the *caveat* of a low CRC case numbers (46 men, 58 in women) in this subgroup.

QCancer-10+LDP consistently provided the best risk prediction. Table 4 shows the sensitivity of the Qancer-10+LDP model in predicting CRC risk over five years across the top 25 centiles of absolute risk. To illustrate, individuals predicted to be in the top 20% of absolute risk by QCancer-10+LDP accounted for 47.6% of male cases and 42.4% of female cases. QCancer-10 and QCancer-10+GWS had lower sensitivity than QCancer-10+LDP (Tables S13-S16). Men in the top 5% of absolute risk by the QCancer-10+LDP model had >3.49-fold increased absolute 5-year risk compared to the median, with a comparable 2.75-fold increase in women. For QCancer-10+GWS this was 3.14-fold and 2.37-fold in men and women respectively, and for QCancer-10 2.37 and 2.06-fold (Table S17). Decision curve analyses confirmed that, across a wide range of probability thresholds, QCancer-10+LDP gave greater net benefit than QCancer-10+GWS and QCancer-10 for both men and women (Figure 3), and predicted a greater number of interventions avoided across clinically relevant thresholds.

**Table 4.** Sensitivity of QCancer-10+LDP models for CRC diagnosis over 5 years of follow-up across the top 25 centiles of absolute risk in males and females. The absolute risk of the top 20% are highlighted.

| Centile | Population per | Absolute 5-year risk | Cases per | Cumulative % cases |
| --- | --- | --- | --- | --- |
| **Males** |  |  |  |  |
| 1 | 1960 | 2.75 | 64 | 3.4 |
| 2 | 1961 | 2.34 | 61 | 6.6 |
| 3 | 1961 | 2.10 | 70 | 10.3 |
| 4 | 1961 | 1.94 | 56 | 13.3 |
| 5 | 1961 | 1.81 | 55 | 16.2 |
| 6 | 1961 | 1.71 | 57 | 19.2 |
| 7 | 1961 | 1.63 | 60 | 22.4 |
| 8 | 1961 | 1.55 | 41 | 24.6 |
| 9 | 1961 | 1.49 | 41 | 26.8 |
| 10 | 1961 | 1.43 | 48 | 29.3 |
| 11 | 1961 | 1.38 | 48 | 31.8 |
| 12 | 1960 | 1.33 | 35 | 33.6 |
| 13 | 1961 | 1.29 | 44 | 35.9 |
| 14 | 1961 | 1.25 | 31 | 37.5 |
| 15 | 1961 | 1.21 | 35 | 39.3 |
| 16 | 1961 | 1.17 | 33 | 41.0 |
| 17 | 1961 | 1.14 | 35 | 42.8 |
| 18 | 1961 | 1.11 | 32 | 44.5 |
| 19 | 1961 | 1.08 | 30 | 46.1 |
| 20 | 1961 | 1.05 | 29 | 47.6 |
| 21 | 1961 | 1.02 | 25 | 48.9 |
| 22 | 1961 | 1.00 | 32 | 50.6 |
| 23 | 1960 | 0.97 | 37 | 52.6 |
| 24 | 1961 | 0.95 | 27 | 54.0 |
| 25 | 1961 | 0.93 | 24 | 55.3 |
| **Females** |  |  |  |  |
| 1 | 2384 | 1.53 | 58 | 4.0 |
| 2 | 2385 | 1.27 | 50 | 7.4 |
| 3 | 2385 | 1.13 | 48 | 10.7 |
| 4 | 2385 | 1.04 | 37 | 13.2 |
| 5 | 2385 | 0.97 | 37 | 15.7 |
| 6 | 2385 | 0.91 | 31 | 17.8 |
| 7 | 2385 | 0.87 | 39 | 20.5 |
| 8 | 2385 | 0.83 | 33 | 22.8 |
| 9 | 2385 | 0.80 | 29 | 24.8 |
| 10 | 2385 | 0.77 | 40 | 27.5 |
| 11 | 2385 | 0.74 | 26 | 29.3 |
| 12 | 2385 | 0.72 | 25 | 31.0 |
| 13 | 2385 | 0.70 | 19 | 32.3 |
| 14 | 2385 | 0.68 | 24 | 33.9 |
| 15 | 2385 | 0.66 | 22 | 35.4 |
| 16 | 2385 | 0.64 | 24 | 37.0 |
| 17 | 2385 | 0.63 | 14 | 38.0 |
| 18 | 2385 | 0.61 | 23 | 39.6 |
| 19 | 2385 | 0.60 | 22 | 41.1 |
| 20 | 2385 | 0.59 | 19 | 42.4 |
| 21 | 2385 | 0.57 | 17 | 43.6 |
| 22 | 2385 | 0.56 | 19 | 44.9 |
| 23 | 2385 | 0.55 | 20 | 46.3 |
| 24 | 2385 | 0.54 | 29 | 48.3 |
| 25 | 2385 | 0.53 | 15 | 49.3 |

**Figure 3.**
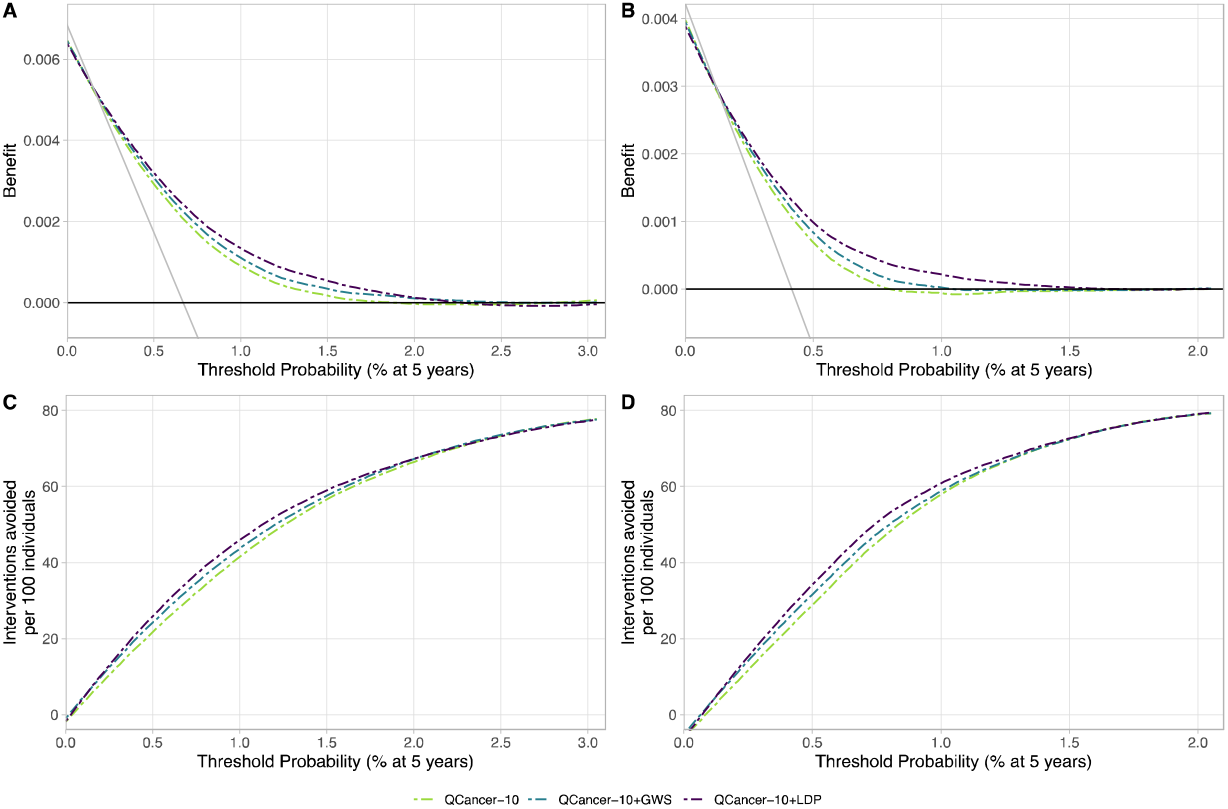
Decision Curve Analysis for QCancer-10+LDP, QCancer-10+GWS, and QCancer-10 models. Figures show net benefit in men (A) and women (B), and interventions avoided per 100 patients tested in men (C) and women (D). The thin grey line in net benefit curves indicates intervention for all, the thick black line no intervention.

By way of illustration, enhanced screening is frequently offered for those with a single first degree relative with CRC (FDRCRC), corresponding to a ∼2.2-fold increased risk. ^27^ QCancer-10+LDP identified 18.2% of men (34.0% of cases) and 7.2% of women (16.5% of cases) as having a relative risk >2.2, of whom 76% and 70% respectively had no FDRCRC (see Table S18 for equivalent values for QCancer-10+GWS and QCancer-10).

## Discussion

We have undertaken the first study to develop and validate new prediction models for colorectal cancer that combine phenotypic risk with genome-wide PRS. We show that combining the non-genetic QCancer-10 model with either genome-wide or GWAS-significant PRS significantly improves model performance and clinical benefit, with greatest improvements seen in the QCancer-10+LDP model. The QCancer-10+LDP models have higher discrimination in UKB than any previously published CRC risk score. ^6,9^

Our models could be used to improve or instigate risk-stratified CRC screening. QCancer-10 ^4^ has recently been recommended to guide shared decision making around CRC screening. ^5^ Our QCancer-10+PRS risk models would provide more accurate risk information for patients and healthcare professionals on which to base screening decisions. Our study predicts that QCancer-10+PRS models have a greater net-benefit and avoid more interventions than QCancer-10 across a wide range of clinically-relevant risk thresholds, with the greatest benefit from QCancer-10+LDP. The sensitivities achieved using QCancer-10+PRS exceed those of other integrated models recently validated in UKBioank. ^6^ The genome-wide SNP genotyping required for LDpred2 is reliably performed from saliva samples, and is rapid, inexpensive and straightforward to analyse. The sensitivities and decision curves provided by QCancer-10+LDP could therefore be used to inform clinical decision making.

Of the PRS methods evaluated, LDpred2-grid and LDpred2-grid-sp models had highest discrimination, explained more of the variation in risk, and were well calibrated. The improvement in performance between the derivation and validation cohorts when using the PRS models probably results from lower genetic homogeneity in the latter. Evaluation of the PRS in a geographically external cohort demonstrates portability of the models. The Geographic Validation Cohort was well matched in age to the derivation cohort, but had a higher proportion of women; prevalence of CRC was higher, at 1.79% compared to 1.51% in the derivation cohort. We would expect performance in Northern European individuals in the general population to be similar to that of the Validation Cohort.

Our PRS findings are in line with a recent study in which a PRS derived using LDpred software (an earlier version of LDpred2) out-performed both machine learning approaches and a 140 GWAS-significant SNP PRS. ^7^ In contrast, a recent study comparing Lassosum software with clumping and thresholding approaches in UK Biobank found the optimal CRC PRS to contain just 87 SNPs, with an age- and sex-adjusted AUC of just 0.617. ^8^ Several genome-wide PRS software tools are now available, with differences in performance across disease types, ^21^ highlighting the importance of evaluating multiple approaches in different phenotypes. Previous studies have found that models combining GWAS-significant PRS and non-genetic risk predictors perform better than PRS alone ^6^ or non-genetic risk factors alone. ^9^ Our work supports and extends this by demonstrating the stepwise improvement in performance obtained with genome-wide PRS.

A key strength of our study is the avoidance of overlap between our GWAS meta-analysis datasets and modelling cohorts, thus reducing overfitting of the PRS and performance optimism. ^10^ We used expected genotype dosages rather than allele counts in each PRS, incorporating uncertainty in genotype imputation, and applied correction for ascertainment bias to effect sizes in the GWAS-significant model. Our GWAS-significant PRS used stringent inclusion criteria, including only SNPs which replicated in our UKB-free meta-analysis.

UKB provides a large sample size, extensive phenotyping, completeness of data recording, and linkage to external datasets. Linkage to cancer registry data in UKB ended in 2015/16 at the time of our study, so we have only been able to follow-up for a median 7 years; updating this will improve risk estimates and permit estimation of risks over a longer follow-up. The UKB age range of ∼40-70 is similar to that of bowel cancer screening (50 to 74 in England and Scotland), but narrower than 25 to 84 used in the original Qcancer-10 study. ^4^ However, model performance in UKB is arguably unlikely to reflect relative performance in the general population, for several reasons. Model performance will vary between populations with different prevalence or risk of a disease – known as the ‘spectrum effect’. As UKB has a lower incidence of disease than the general population of screening age, one might expect sensitivity to increase (which is of benefit in a screening test) when applied to a population with higher risk. ^28^ Furthermore, all of our models appeared to perform less well in females. For PRS, wide confidence intervals in the Geographic Validation Cohort mean this finding should be interpreted with caution, but for models that include QCancer-10, this difference was not unexpected. The known healthy volunteer bias that exists in UKB is especially marked in women (for example, the reduction in all-cause mortality and overall cancer incidence in UKB relative to the general population is greater for women than men). ^29^ In addition, the available sample size and number of incident cases for women in our Integrated Modelling Cohort fell slightly short of sample size requirements (see Supplementary Methods). As a result our estimates of risk may be less precise for women, and external validation would be essential prior to implementation. The QCancer-10 model has previously been shown to perform worse when validated in UKB than in the QResearch validation. ^4^ This is likely to be due to the differences in age distribution between the general population sample used to develop the original QCancer-10 score and the more restricted UKB sample in this study. External validation of a separate QCancer (colorectal) score for symptomatic patients in an independent population-based cohort showed comparable performance to the discovery study. ^30^ Overall, risk model performance should be validated in a population representative of the screening population, and we have shown that PRS calibration can be largely corrected in new (ethnically similar) populations by recalibration.

Further limitations of our study may include unknown differences in the demographics of the contributing base GWAS datasets and UKB. We did not include Mendelian CRC syndromes in the genetic model, and doing so would almost certainly improve risk prediction. Another major limitation of our study, and PRS generally, is that most models are developed in individuals of European ethnicity. Although most CRC risk SNPs appear to be shared across ethnic groups, quantitative risk estimates cannot readily be transferred across populations, ^31^ and, as anticipated, our PRS performed poorly in the Minority Ethnic Validation Cohort. ^31-33^ As minority ethnic populations often have higher CRC associated mortality and lower screening uptake, ^34^ further work is urgently needed to expand PRS for CRC in these populations to avoid exacerbating existing health inequalities.

With a renewed focus on prevention and early diagnosis in healthcare, ^35^ refined risk prediction is likely to have a significant role. It has been proposed that risk scores could be used to improve diagnosis in symptomatic individuals (for example, https://ourfuturehealth.org.uk/). However, the combined risk score alone is unlikely to achieve the performance standards required for diagnostic tests, and would need to be evaluated carefully in combination with quantitative faecal immunochemical test (FIT) results, symptoms and clinical signs.

In population cancer screening programmes, in contrast, a risk score with moderate predictive value has considerable potential for improving current performance through patient stratification. Our risk score predicts that ∼10% of the population aged ∼40-70 have relative risks of CRC high enough to warrant enhanced surveillance under guidelines used in familial risk. ^36^ The risk models constructed here perform at a level that may well be clinically useful in population screening. ^37^ In practice, we envisage that risk scores are likely to be used alongside FIT to allocate colonoscopy more effectively, maintaining universal access to screening whilst improving performance. Nevertheless, we emphasise that validation of the risk score in a cohort representative of the screening population and evaluation in a prospective trial are required to evaluate the performance, acceptability and cost effectiveness of a combined risk model in a FIT-based bowel cancer screening programme.

## Data Availability

UK Biobank data can be obtained through http://www.ukbiobank.ac.uk/. Genotype data are available in the European Genome-phenome Archive under accession numbers EGAS00001005412, EGAS00001005421, or from the Edinburgh University DataShare Repository (https://datashare.ed.ac.uk/). Finnish cohort samples can be requested from the THL Biobank https://thl.fi/en/web/thl-biobank. PRS SNP inclusion lists and model specifications will be deposited in the PGS catalogue repository (https://www.pgscatalog.org/). Risk scores for UKB participants will be returned to UK Biobank for use by approved researchers.

https://datashare.ed.ac.uk/

https://thl.fi/en/web/thl-biobank

## Contributors

All authors contributed to study conception and design, with development of PRS and statistical analysis led by SEWB, IT and JHC. IT, MD and RH provided data. SEWB, IT, PL and RH have accessed and verified the underlying data. SEWB carried out primary data analysis. SEWB completed the statistical analysis under supervision of IT and JHC. IT, JCH, SE and JEE supervised the project. SEWB and IT wrote the first draft of the manuscript. All authors contributed to critical revision and editing of the manuscript, and have approved the final version. JHC and IT are guarantors.

## Ethics Approval

This research has been conducted using data from UK Biobank, a major biomedical database, http://www.ukbiobank.ac.uk/. The UK Biobank study has ethical approval from the North West Multi-centre Research Ethics Committee (16/NW/0274). This study was performed under UK Biobank application number 8508. All contributing GWAS studies were undertaken with ethical review board approval at respective study centres as detailed in Law *et al*.^*16*^

## Funding/acknowledgements

We thank all individuals who agreed to participate in the contributing GWAS studies and in UK Biobank, and the investigators, research associates and wider teams involved in these studies. We thank the authors of LDpred2 for their instructive PRS tutorial and publicly available code.

SB is supported by an MRC Clinical Research Training Fellowship (MR/P001106/1). JEE and SW receive funding from the Oxford NIHR Biomedical Research Centre (BRC). This work of the Houlston Laboratory (PL, RH) is supported by a grant from Cancer Research UK (CRUK) (C1298/A25514). JHC received funding from the John Fell Oxford University Press Research Fund, grants from CRUK grant number C5255/A18085, through the CRUK Oxford Centre, grants from the Oxford Wellcome Institutional Strategic Support Fund (204826/Z/16/Z) and other research councils, during the conduct of the study. MD is funded by CRUK Programme Grant C348/A12076. IT is funded by CR-UK Programme Grant C6199/A27327. The research was supported by the Wellcome Trust Core Award Grant Number 203141/Z/16/Z with funding from the NIHR Oxford BRC. The views expressed are those of the author(s) and not necessarily those of the NHS, the NIHR or the Department of Health. Funding bodies had no role in the design, analysis, writing or decision to submit.

## Transparency

The lead author (IT) affirms that the manuscript is an honest, accurate, and transparent account of the study being reported; that no important aspects of the study have been omitted; that discrepancies from the study as planned have been explained, and that the paper conforms to transparency policy of the International Committee of Medical Journal Editors uniform requirement for manuscripts submitted to biomedical journals.

## References

1. Keum N, Giovannucci E. Global burden of colorectal cancer: emerging trends, risk factors and prevention strategies. Nat Rev Gastroenterol Hepatol. 2019;16(12):713–32.DOI: 10.1038/s41575-019-0189-8

2. Pashayan N, Morris S, Gilbert FJ, Pharoah PDP. Cost-effectiveness and Benefit-to-Harm Ratio of Risk-Stratified Screening for Breast Cancer: A Life-Table Model. JAMA Oncol. 2018;4(11):1504–10.DOI: 10.1001/jamaoncol.2018.1901

3. Usher-Smith JA, Harshfield A, Saunders CL, et al. External validation of risk prediction models for incident colorectal cancer using UK Biobank. Br J Cancer. 2018;118(5):750–9.DOI: 10.1038/bjc.2017.463

4. Hippisley-Cox J, Coupland C. Development and validation of risk prediction algorithms to estimate future risk of common cancers in men and women: prospective cohort study. BMJ Open. 2015;5(3):e007825.DOI: 10.1136/bmjopen-2015-007825

5. Helsingen LM, Vandvik PO, Jodal HC, et al. Colorectal cancer screening with faecal immunochemical testing, sigmoidoscopy or colonoscopy: a clinical practice guideline. BMJ. 2019;367:l5515.DOI: 10.1136/bmj.l5515

6. Saunders CL, Kilian B, Thompson DJ, et al. External Validation of Risk Prediction Models Incorporating Common Genetic Variants for Incident Colorectal Cancer Using UK Biobank. Cancer Prev Res (Phila). 2020;13(6):509–20.DOI: 10.1158/1940-6207.CAPR-19-0521

7. Thomas M, Sakoda LC, Hoffmeister M, et al. Genome-wide Modeling of Polygenic Risk Score in Colorectal Cancer Risk. Am J Hum Genet. 2020;107(3):432–44.DOI: 10.1016/j.ajhg.2020.07.006

8. Fritsche LG, Patil S, Beesley LJ, et al. Cancer PRSweb: An Online Repository with Polygenic Risk Scores for Major Cancer Traits and Their Evaluation in Two Independent Biobanks. Am J Hum Genet. 2020;107(5):815–36.DOI: 10.1016/j.ajhg.2020.08.025

9. Kachuri L, Graff RE, Smith-Byrne K, et al. Pan-cancer analysis demonstrates that integrating polygenic risk scores with modifiable risk factors improves risk prediction. Nat Commun. 2020;11(1):6084.DOI: 10.1038/s41467-020-19600-4

10. Wray NR, Yang J, Hayes BJ, Price AL, Goddard ME, Visscher PM. Pitfalls of predicting complex traits from SNPs. Nat Rev Genet. 2013;14(7):507–15.DOI: 10.1038/nrg3457

11. Collins GS, Reitsma JB, Altman DG, Moons KG. Transparent Reporting of a multivariable prediction model for Individual Prognosis Or Diagnosis (TRIPOD). Ann Intern Med. 2015;162(10):735–6.DOI: 10.7326/L15-5093-2

12. Wand H, Lambert SA, Tamburro C, et al. Improving reporting standards for polygenic scores in risk prediction studies. Nature. 2021;591(7849):211–9.DOI: 10.1038/s41586-021-03243-6

13. Bycroft C, Freeman C, Petkova D, et al. The UK Biobank resource with deep phenotyping and genomic data. Nature. 2018;562(7726):203–9.DOI: 10.1038/s41586-018-0579-z

14. Sudlow C, Gallacher J, Allen N, et al. UK biobank: an open access resource for identifying the causes of a wide range of complex diseases of middle and old age. PLoS Med. 2015;12(3):e1001779.DOI: 10.1371/journal.pmed.1001779

15. Office for National Statistics. Cancer registration statistics, England [Available from: https://www.ons.gov.uk/peoplepopulationandcommunity/healthandsocialcare/conditionsanddiseases/datasets/cancerregistrationstatisticscancerregistrationstatisticsengland. Accessed September 2020].

16. Law PJ, Timofeeva M, Fernandez-Rozadilla C, et al. Association analyses identify 31 new risk loci for colorectal cancer susceptibility. Nat Commun. 2019;10(1):2154.DOI: 10.1038/s41467-019-09775-w

17. Liu JZ, Tozzi F, Waterworth DM, et al. Meta-analysis and imputation refines the association of 15q25 with smoking quantity. Nat Genet. 2010;42(5):436–40.DOI: 10.1038/ng.572

18. Huyghe JR, Bien SA, Harrison TA, et al. Discovery of common and rare genetic risk variants for colorectal cancer. Nat Genet. 2019;51(1):76–87.DOI: 10.1038/s41588-018-0286-6

19. Bigdeli TB, Lee D, Webb BT, et al. A simple yet accurate correction for winner’s curse can predict signals discovered in much larger genome scans. Bioinformatics. 2016;32(17):2598–603.DOI: 10.1093/bioinformatics/btw303

20. Prive F, Vilhjalmsson BJ, Aschard H, Blum MGB. Making the Most of Clumping and Thresholding for Polygenic Scores. Am J Hum Genet. 2019;105(6):1213–21.DOI: 10.1016/j.ajhg.2019.11.001

21. Prive F, Arbel J, Vilhjalmsson BJ. LDpred2: better, faster, stronger. Bioinformatics. 2020;36(22-23):5424–31.DOI: 10.1093/bioinformatics/btaa1029

22. Royston P, Altman DG. External validation of a Cox prognostic model: principles and methods. BMC Med Res Methodol. 2013;13:33.DOI: 10.1186/1471-2288-13-33

23. Steyerberg EW, Vickers AJ, Cook NR, et al. Assessing the Performance of Prediction Models A Framework for Traditional and Novel Measures. Epidemiology. 2010;21(1):128–38.DOI: 10.1097/EDE.0b013e3181c30fb2

24. Harrell FE. Regression Modeling Strategies: with Applications to Linear Models, Logistic Regression, and Survival Analysis. New York, NY: Springer; 2001.

25. Riley RD, Ensor J, Snell KIE, et al. Calculating the sample size required for developing a clinical prediction model. BMJ. 2020;368.DOI: ARTN m441 10.1136/bmj.m441

26. R Core Team. R: A language and environment for statistical computing. 2019 https://www.R-project.org/.

27. Johns LE, Houlston RS. A systematic review and meta-analysis of familial colorectal cancer risk. Am J Gastroenterol. 2001;96(10):2992–3003.DOI: 10.1111/j.1572-0241.2001.04677.x

28. Usher-Smith JA, Sharp SJ, Griffin SJ. The spectrum effect in tests for risk prediction, screening, and diagnosis. BMJ. 2016;353:i3139.DOI: 10.1136/bmj.i3139

29. Fry A, Littlejohns TJ, Sudlow C, et al. Comparison of Sociodemographic and Health-Related Characteristics of UK Biobank Participants With Those of the General Population. Am J Epidemiol. 2017;186(9):1026–34.DOI: 10.1093/aje/kwx246

30. Collins GS, Altman DG. Identifying patients with undetected colorectal cancer: an independent validation of QCancer (Colorectal). Br J Cancer. 2012;107(2):260–5.DOI: 10.1038/bjc.2012.266

31. Martin AR, Kanai M, Kamatani Y, Okada Y, Neale BM, Daly MJ. Clinical use of current polygenic risk scores may exacerbate health disparities. Nat Genet. 2019;51(4):584–91.DOI: 10.1038/s41588-019-0379-x

32. Campbell C, Douglas A, Williams L, et al. Are there ethnic and religious variations in uptake of bowel cancer screening? A retrospective cohort study among 1.7 million people in Scotland. BMJ Open. 2020;10(10):e037011.DOI: 10.1136/bmjopen-2020-037011

33. Hirst Y, Stoffel S, Baio G, McGregor L, von Wagner C. Uptake of the English Bowel (Colorectal) Cancer Screening Programme: an update 5 years after the full roll-out. Eur J Cancer. 2018;103:267–73.DOI: 10.1016/j.ejca.2018.07.135

34. Carethers JM, Doubeni CA. Causes of Socioeconomic Disparities in Colorectal Cancer and Intervention Framework and Strategies. Gastroenterology. 2020;158(2):354–67.DOI: 10.1053/j.gastro.2019.10.029

35. NHS England. The NHS long term plan 2019 [Available from: https://www.longtermplan.nhs.uk/.

36. Monahan KJ, Bradshaw N, Dolwani S, et al. Guidelines for the management of hereditary colorectal cancer from the British Society of Gastroenterology (BSG)/Association of Coloproctology of Great Britain and Ireland (ACPGBI)/United Kingdom Cancer Genetics Group (UKCGG). Gut. 2020;69(3):411–44.DOI: 10.1136/gutjnl-2019-319915

37. Sud A, Turnbull C, Houlston R. Will polygenic risk scores for cancer ever be clinically useful? NPJ Precis Oncol. 2021;5(1):40.DOI: 10.1038/s41698-021-00176-1

