## Supplementary Data for "Integrating genome-wide polygenic risk scores and non-genetic risk to predict colorectal cancer diagnosis: a cohort study in UK Biobank"

**Table S8. Apparent, internally and externally validated polygenic risk score (PRS) performance in Cox's proportional hazards models (adjusting for age, sex, array and first 4 principal components).** 20

**Figure S9. Kaplan-Meier curves across four risk groups (group 4 being highest risk) for PRS in the Geographic Validation Cohort compared with the Test Cohort** 21

**Figure S10. Calibration plots of PRS models in Cox models in the Geographic Validation Cohort** 22

**Table S9. Subgroup analysis of PRS Cox model performance by sex** 23

**Figure S11. Calibration of PRS in Cox models by sex in the Geographic Validation Cohort.** 24

**Figure S12. Observed and predicted probabilities of CRC for PRS Cox models across 5 year age bands in the Geographic Validation Cohort** 25

**Figure S13. Kaplan-Meier curves across four risk groups (group 4 being highest risk) for PRS in the Minority Ethnic Validation Cohort compared with the Test Cohort** 26

**Figure S14. Calibration plots of PRS models in Cox model in the Minority Ethnic Validation Cohort.** 27

**Table S10. Characteristics of the UKB Integrated Modelling Cohort used for QCancer-10 validation, compared with the QCancer-10 derivation cohort.** 28

**Figure S15. Calibration of QCancer-10 over 5-8 years of follow-up.** 29

**Table S11. Expected/observed ratio of risk over 5-8 years of follow-up for male and female for QCancer-10+LDP, QCancer-10+GWS and QCancer-10 models in subgroups analyses.** 30

**Figure S16. Observed and predicted probabilities of CRC by age for male and female for QCancer-10+LDP, QCancer-10+GWS and QCancer-10 models.** 31

**Figure S17. Calibration plots for individuals with a first-degree family history of CRC in QCancer-10+PRS and QCancer-10 models.** 32

**QCancer-10+PRS model specification** 33

**Table S12. Interaction terms in QCancer-10+LDP and QCancer-10+GWS models.** 33

**Figure S18. Marginal effect of QCancer-10 risk score in interaction with PRS in male QCancer-10+LDP and QCancer-10+GWS models** 33

**Table S13. Sensitivity of QCancer-10+GWS models for CRC diagnosis over 5 years of follow-up across top 25 centiles of absolute risk** 35

**Table S14. Sensitivity of QCancer-10 for CRC diagnosis over 5 years of follow-up across top 25 centiles of absolute risk** 36

**Table S15. Sensitivity of QCancer-10+LDP across top 25 centiles of relative risk.** 37

**Table S16. Sensitivity of QCancer-10+GWS across top 25 centiles of relative risk.** 38

**Table S17. Fold-increase in absolute risk between 95$^{th}$ centile and median risk for QCancer-10+LDP, QCancer-10+GWS and QCancer-10 models** 39

**Table S18. Percentage of population and cases with relative risk > 2** 39

*References* 40

### Supplementary Methods

#### Base Genome-wide Association Study Meta-analysis

The base dataset for polygenic risk score (PRS) development was obtained through meta-analysis of the datasets included in Law *et al.*,[1] excluding the UK Biobank dataset. Summary data from the following genome-wide association study (GWAS) datasets was therefore included: NSCCG-OncoArray; SCOT; SOCCS/GS; SOCCS/LBC; CCFR1; CCFR2; COIN; CORSA; Croatia; DACHS; FIN; UK1; Scotland1; VQ58. The contributing datasets, genotyping and imputation information, quality control (QC) and study approvals are described in detail in Law *et al.*[1]

#### Cancer Incidence Calculation

Whole UKB cohort CRC incidence rates were calculated based on linked registry cases, without removal of prevalent cases, to reflect registration as would occur in national data. In addition ASIRs were calculated in the Integrated Modelling Cohort, in which prevalent cases were removed and cases identified through cancer and death registry, and linked hospital inpatient data; follow-up duration was as defined in the main methods. This analysis used R packages 'survival' and 'epitools'.[2,3]

#### PRS Sample QC and dataset definitions

We performed standard per-person QC on all individuals with imputed genetic data available, removing those with sex chromosome aneuploidy, sex-mismatch and an excess of relatives in the dataset. The Derivation Cohort (see Figure 1) included individuals identified by UKB as having white-British ancestry (on the basis of self-report and principal components analysis), and recruited through English and Welsh centres. We performed further QC on this cohort,[4] removing those who were not included in the PCA calculation (mainly due to relatedness), and restricting further to a genetically homogeneous subset (those within log-distance of 5 following computation of a robust Mahalanobis distance), resulting in a dataset of 310664 individuals.

The Geographic Validation Cohort comprised 34152 individuals recruited in Scotland and of European ancestry (UK Biobank self-reported ethnicities of "British", "Irish", "White", and "Any other white background") passing standard QC. Scotland was chosen for validation as this cohort contained more than the recommended number of cases for model validation (a minimum of 100, and ideally 200, cases),[5] and represents a population with different demographics to England and Wales, testing the models portability.

A Minority Ethnic Validation Cohort (n = 27503) comprised all UK Biobank participants passing standard QC with self-reported ethnicities not in the above categories (including individuals who responded "Do not know" and "Prefer not to answer", but not those with missing ethnicity data).

Ten thousand randomly selected individuals were used as a subset in which the linkage disequilibrium (LD) matrix was computed (used for C+T, SCT and LDpred2 models).[4] However, owing to the relatively small number of cases in this set (136), we added an additional randomly selected 20000 individuals to derive a Training Cohort of 30000 individuals for PRS hyper-parameter selection. The remaining 280664 individuals comprised the Test Cohort in which PRS performance was evaluated.

We used imputed dosage data from UK Biobank, restricting variants to those included in HapMap3, and with matched SNPs in the base data. Of 12972739 SNPs present in the base GWAS summary statistics, 1798524 ambiguous SNPs were removed and 1117002 variants matched with UK Biobank data. QC was performed as recommended by Privé *et al.*[4] on the summary statistics, comparing standard deviations of genotypes in the summary statistics ($SD_{ss}$) and 10000 individuals from the LDpred2 Training Cohort ($SD_{ldtr}$), and removing variants where $SD_{ss} < 0.5(SD_{ldtr})$, $SD_{ss} > 0.1 + SD_{ldtr}$, $SD_{ss} < 0.1$, or $SD_{ldtr} < 0.05$ (Figure S1),[4] leaving 1104409 SNPs included in analysis. Following QC, the minimum INFO score was 0.411.

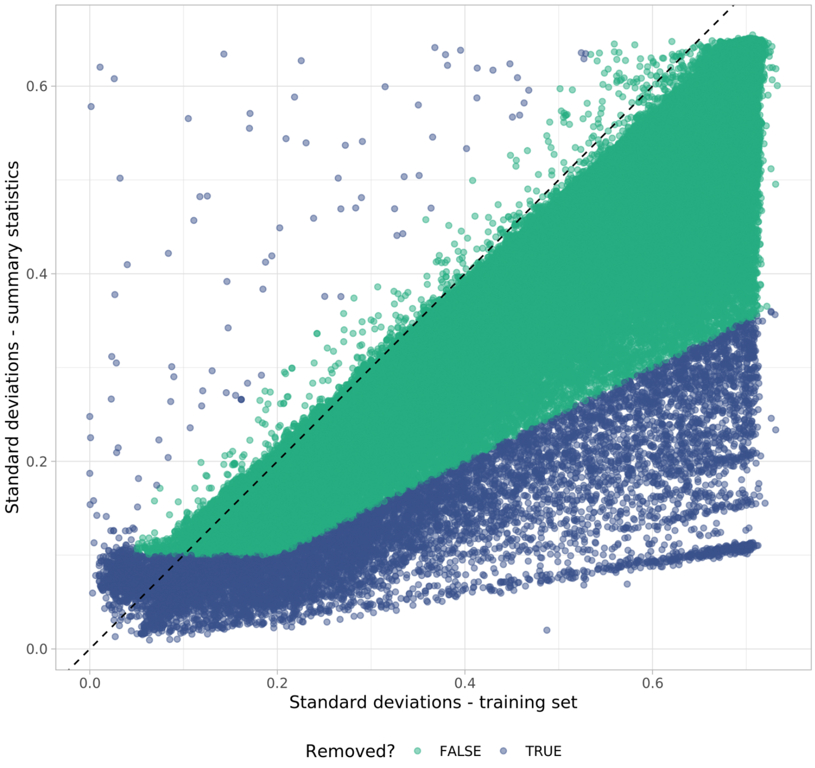

**Figure S1. SNP QC based on standard deviations of genotypes in LDpred2 ‘validation’ dataset and base data summary statistics (after Privé et al.)**

**GWAS significant PRS**

We manually curated a list of SNPs derived from previously published GWAS in European populations including Law *et al.* and Huyghe *et al.*[1,6] and the references within these. We excluded SNPs which did not reach genome-wide significance ($p<5x10^{-8}$) in our base meta-analysis, and used the effect sizes from our meta-analysis, adjusted for the winner's curse using the False Discovery Rate Inverse Quantile Transformation (FIQT) method.[7] Where SNPs were reported at the same loci in different studies and were correlated at $r^2>0.1$ we retained the most significantly associated SNP. We confirmed that all of the included SNPs imputed well in the UKB data with INFO scores > 0.9. The PRS was calculated as the sum of allele dosages weighted by their effect sizes.

**C+T and SCT PRS**

Clumping and thresholding approaches to SNP selection generate PRS scores across a range of LD $r^2$ values (with a given window size for clumping selected) and association p value thresholds. We used R package *bigsnpr* by Privé et al.[8] to generate scores across a grid of $r^2$, p-value threshold, and clumping window size values. Default parameters were used: clumping $r^2$of 0.01, 0.05, 0.1, 0.2, 0.5, 0.8, 0.95; 50 p-value thresholds spaced equally between 0.1 and the most significant p-value on the log scale; and a base clumping window size of 50, 100, 200 and 500 (where actual window size in kb is the base size divided by clumping $r^2$).

From this grid, a maximum score was selected based on AUC (the C+T score in this paper), and stacking used to learn the optimal linear combination of scores generated through efficient penalised regression (the SCT score).[8]

**LDpred2 PRS**

LDpred2[4] uses a Bayesian approach to SNP selection and shrinkage for PRS, based on an LD matrix and GWAS summary statistics, implemented in the R package bigsnpr. This updated version of LDpred has been demonstrated to provide higher predictive performance, particularly with large GWAS sample size as in this study,[4] and also addresses previous instability issues.[9] The use of a larger window of 3cM (using genetic distance rather than number of bases) improves performance when causal variants are located in regions of long-range LD, such as HLA regions. Colorectal cancer-associated variants in these regions have recently been reported,[1,6] and this improvement may therefore be of benefit in CRC-prediction. LDpred2 also evaluates more hyper-parameters (a grid of 126 instead of 7 in LDpred).

There are multiple options for PRS construction within LDpred2. An infinitesimal model (LDpred2-inf), in which all makers are assumed to be causal; grid models (LDpred2-grid) in the hyper-parameters SNP heritability, $h^2$, proportion of causal variants, *p,* and optionally sparsity, are tuned in a validation set; and an auto model (LDPred2-auto) in which sparsity and SNP heritability are estimated automatically, negating the need for a validation set. LDpred2 estimates heritability calculated from constrained LD score regression. The estimate for this dataset was 0.1602065.

We evaluated LDpred2-inf and LDpred2-grid models (sparse and non-sparse), running them genome-wide as recommended. LDpred2-grid outputs SNP effect sizes for each of the grid values; the optimally performing model was then selected based on best Z-score for the logistic regression slope (Figure S2), in which we adjusted for array platform and first 4 principal components (PCs).

Clumping and thresholding and LDpred2 modelling code was adapted from code provided by Privé *et al.* at https://github.com/Privéfl/paper-ldpred2/tree/master/code, and their accompanying LDpred2 tutorial.[4]

**Evaluation of polygenic risk score performance**

Each PRS was evaluated in logistic regression and Cox models, adjusting for age, sex, array and 4 principal components. Age, sex and PCs were all modelled as continuous variables, assuming a linear relationship. For Cox models we confirmed proportional hazards assumptions held through visual inspection of plots of Schoenfeld residuals. We evaluated potential interactions between PRS and age by examining the prognostic strength and significance of interaction terms based on Wald $\chi^2$statistics, and plotting marginal effects of PRS with age. We compared model performance to a reference model, containing age, sex, array and 4 principal components, to assess the contribution of the PRS to model performance.

Further models were also derived which did not adjust for age and sex,[10] to evaluate the contribution which these factors (known to be independent predictors for CRC risk) made to the performance of the full model.

In order to compare PRS distributions for each cohort, and effect sizes per SD of each PRS, we standardised the PRS to have a mean of 0 and standard deviation of 1 in the Test Cohort. We also used these standardised scores in plots of marginal effects of PRS in interaction with age. Remaining analyses used non-standardised scores.

**Validation of QCancer-10**

Validation of QCancer-10 in UKB permits evaluation of model performance in a population of approximately bowel screening age. Full QCancer-10 model specification is available at https://www.qcancer.org/15yr/colorectal/.[11]

CRC outcomes were identified as described in the main paper. Of note, in QCancer-10 (colorectal cancer) development ICD-10 codes for anal cancer were included in case definition. We did not include these in this study, as anal cancers are of a different aetiology to CRC, and bowel cancer screening does not aim to detect these lesions. Previous medical history, alcohol and smoking status, and family history were all taken from self-reported data in baseline touch-screen and verbal UKB assessment centre interviews.

Mapping of ethnicity, smoking and alcohol intake is given in Table S1. Ethnicity was coded from self-reported ethnicity (UKB field 21000). Smoking history was compiled from the smoking summary field (field 20116), frequency of smoking (field 1239) and number of cigarettes smoked (field 3456). To calculate alcohol intake, reported alcohol intake frequency (field 1558) was combined with detailed drink-based intake reported in glasses/pints at touchscreen interview. Drinks intake was converted to units using NHS Choices Livewell alcohol units (as in Usher-Smith *et al.*[12]), and average daily units calculated.

Previous medical history of cancers was taken from self-reported cancer and non-cancer illnesses (fields 20001 and 20002) at touch-screen interview (Table S2).

Family history in UKB is for first degree relatives, detailed for father, mother and siblings separately; we considered positive family history to be CRC in any of these relatives. In QCancer-10 development, absence of data carries the assumption that the individual does not have any family history; family history was therefore coded as missing only if the answer for all of these was either 'Do not know' or 'Prefer not to answer'.

Distributions of continuous predictors were evaluated. One implausible value for BMI was set to missing and otherwise all values were retained. Of note there are a very small number of UKB participants aged 38-39 and 71-73 years at baseline assessment, who were included in our modelling.

**Table S1: Mapping of UK Biobank ethnicity, smoking and alcohol data to QCancer-10 coding**

| QCancer-10 Coding | UK Biobank Coding |
|---|---|
| **Ethnicity** | |
| White/not recorded | White, British, Irish, Any other white background, Prefer not to answer, Do not know, Missing |
| Indian | Indian |
| Pakistani | Pakistani |
| Bangladeshi | Bangladeshi |
| Other Asian | Asian or Asian British, Any other Asian background |
| Caribbean | Caribbean |
| Black African | African |
| Chinese | Chinese |
| Other | Black or Black British, Any other Black background, Mixed, White and Black Caribbean, White and Black African, White and Asian, Any other mixed background, Other ethnic group |
| **Smoking** | |
| Non-smoker | Smoking summary = 'Never' |
| Ex-smoker | Smoking summary = 'Previous' |
| Light smoker | Smoking summary = 'Current' AND Cigarettes < 10 OR frequency = 'Only occasionally' |
| Moderate smoker | Smoking summary = 'Current' AND Cigarettes = 10-19 |
| Heavy smoker | Smoking summary = 'Current' AND Cigarettes >20 |
| Missing | Smoking summary = 'Missing'/ 'Prefer not to answer' |
| **Alcohol** | |
| Non-drinker | Alcohol frequency = 'Never' |
| Trivial drinker | <1 calculated daily unit |
| Light drinker | 1-2 calculated daily units |
| Moderate drinker | 3-6 calculated daily units |
| Heavy drinker | 7-9 calculated daily units |
| Very heavy drinker | 10 or more calculated daily units |
| Missing | Alcohol frequency = 'Missing'/ 'Prefer not to answer' |

**Table S2: UK Biobank codes self-reported medical history for QCancer-10 predictors**

| Medical condition | UK Biobank codes |
|---|---|
| Diabetes | 1223, 1220 |
| Ulcerative colitis | 1463 |
| Bowel polyps | 1460 |
| Breast cancer | 1002 |
| Uterine cancer | 1040 |
| Ovarian cancer | 1039 |
| Cervical cancer | 1041 |
| Lung cancer | 1001 |
| Blood cancers | 1047, 1048, 1050, 1051, 1052, 1053, 1055, 1056, 1058 |
| Oral cancers | 1004, 1005, 1006, 1010, 1011, 1012, 1015, 1077, 1078, 1079 |

### Integrated model development

Riley *et al.* propose minimum sample size requirements for developing new prediction models which go beyond the historically recommended 20 events per variable, implemented in R package pmsampsize.[13] This uses the anticipated Cox-Snell $R^2$, number of predictors considered in the model, duration of follow-up, and expected event rate to calculate sample size and number of cases required.

We derived the Cox-Snell $R^2$ as described by Riley *et al.*[13] from the C-statistics from the open cohort of QCancer-10 validation performed in UK Biobank by Usher-Smith *et al.*[12] (0.70 and 0.65 for male and female models respectively), and mean follow-up and CRC rates calculated for individuals available for the Integrated Modelling Cohort. The number of predictors included in the integrated model for each sex was calculated as follows for QCancer-10 risk score components: 1 for each degree of freedom of each categorical variable (alcohol intake = 5; ethnicity = 8; smoking = 4); 1 each for continuous variables (BMI, Townsend Deprivation Score); 1 for each boolean predictor; 1 parameter for each fractional polynomial term for age; and 2 parameters for each interaction term calculated; 1 for the QCancer-10 risk score itself. With 1 additional parameter added for the PRS, this totalled 34 parameters for men and 33 for women.

Sample size calculations indicated that for the integrated male model, 27.43 events per candidate predictor parameter (EPP) are needed, giving a minimum sample size of 94996 and 933 events. As a result of lower CRC incidence and expected model performance in women, the EPP required was 47.53, minimum sample size 253780, with 1569 events. Whilst the numbers required for the male model are readily achievable, the sample size and cases available for the female model fall short in the our available Integrated Modelling Cohort (n = 238496, including 1458 cases). Whilst we continued with model development, for the female integrated models the estimate of outcome risk may be less precise, and the model may be more subject to over-fitting.[14] External validation of the integrated model will be essential prior to implementation.

We constructed Cox models in the Integrated Modelling Cohort including two predictors: the risk score from QCancer-10 and a PRS. We developed male and female models separately, and compared the use of the top-performing genome-wide PRS, and the GWAS-sig PRS. We truncated the lower and upper 0.5% of the distributions of each predictor to the outer bounds.[15] Inspection of Schoenfeld residuals showed that the proportional hazard assumption held. We evaluated the use of multiple fractional polynomials to model the predictors. We assessed possible interactions between the predictors by visual inspection of plots of marginal effects of the QCancer-10 risk score across PRS values, and examining the prognostic strength and significance of interaction terms based on Wald $\chi^2$statistics.

### Software

R package bigsnpr v1.5.2[16] was used for genome-wide PRS development, epitools v 0.5-10.1,[2] rms v5.1-4,[17] mfp v1.5 .2,[18] and survival v3.1-8[3] for modelling, and packages from the tidyverse suite[19] for data analysis and presentation.

### Supplementary Results

**Table S3: Characteristics and missingness of predictor values for the whole UKB cohort, excluding individuals with prevalent CRC**. Values are numbers (%) unless otherwise indicated. CRC – colorectal cancer, IQR – interquartile range, NA – not applicable. *not included in model for females but provided for information.

| | Male | Female |
|---|---|---|
| Age (years), median (IQR) | 58.0 (14.0) | 57.0 (13.0) |
| Ethnicity | | |
| White/not recorded | 215121 (94.6) | 257402 (94.6) |
| Indian | 3003 (1.3) | 2933 (1.1) |
| Pakistani | 1118 (0.5) | 716 (0.3) |
| Bangladeshi | 159 (0.1) | 74 (0.0) |
| Other Asian | 996 (0.4) | 857 (0.3) |
| Caribbean | 1637 (0.7) | 2855 (1.0) |
| Black African | 1701 (0.7) | 1677 (0.6) |
| Chinese | 581 (0.3) | 989 (0.4) |
| Other | 3107 (1.4) | 4529 (1.7) |
| Follow-up (years), median (IQR) | 7.08 (1.34) | 7.09 (1.31) |
| < 5 years | 405 (0.2) | 490 (0.2) |
| Townsend deprivation index, median (IQR) | -2.1 (4.3) | -2.1 (4.1)* |
| Missing | 293 (0.1) | 325 (0.1) |
| BMI (kg/m2), median (IQR) | 27.3 (5.1) | 26.1 (6.3)* |
| Missing | 1639 (0.7) | 1448 (0.5) |
| Smoking status | | |
| Non-smoker | 110840 (48.7) | 161336 (59.3) |
| Ex-smoker | 86721 (38.1) | 84928 (31.2) |
| Light smoker | 11009 (4.8) | 10158 (3.7) |
| Moderate smoker | 6958 (3.1) | 8402 (3.1) |
| Heavy smoker | 10474 (4.6) | 5708 (2.1) |
| Missing | 1421 (0.6) | 1500 (0.6) |
| Alcohol intake | | |
| Non-drinker | 14472 (6.4) | 25889 (9.5) |
| Trivial drinker | 48955 (21.5) | 109497 (40.3) |
| Light drinker | 66332 (29.2) | 87095 (32) |
| Moderate drinker | 69467 (30.5) | 42795 (15.7) |
| Heavy drinker | 17115 (7.5) | 4374 (1.6) |
| Very heavy drinker | 10323 (4.5) | 1646 (0.6) |
| Missing | 759 (0.3) | 736 (0.3) |
| Family history of CRC | 21638 (9.5) | 24773 (9.1) |
| Missing | 9652 (4.2) | 7262 (2.7) |
| Diabetes | 15513 (6.8) | 9294 (3.4) |
| Missing | 358 (0.2) | 298 (0.1) |
| Colorectal polyps | 711 (0.3) | 708 (0.3) |
| Missing | 378 (0.2) | 310 (0.1) |
| Ulcerative colitis | 1187 (0.5) | 1379 (0.5) |
| Missing | 378 (0.2) | 310 (0.1) |
| Breast cancer | NA | 11165 (4.1) |
| Missing | NA | 649 (0.2) |

| | | |
|---|---|---|
| Uterine cancer | NA | 1194 (0.4) |
| Missing | NA | 649 (0.2) |
| Ovarian cancer | NA | 811 (0.3) |
| Missing | NA | 649 (0.2) |
| Cervical cancer | NA | 1985 (0.7) |
| Missing | NA | 649 (0.2) |
| Lung cancer | 149 (0.1) | NA |
| Missing | 631 (0.3) | NA |
| Blood cancer | 1356 (0.6) | NA |
| Missing | 631 (0.3) | NA |
| Oral cancer | 576 (0.3) | NA |
| Missing | 631 (0.3) | NA |
| Imputed genetic data passing standard QC | 220923 (97.1) | 262403 (96.5) |
| Missing | 6500 (2.9) | 9629 (3.5) |

**Table S4: Demographics of derivation and validation cohorts used in PRS development (logistic regression modelling cohorts)**

| | Derivation Training | | Derivation Test | | Geographic Validation | | Minority Ethnic Validation | |
|---|---|---|---|---|---|---|---|---|
| | **Controls (n = 29554)** | **Cases (n = 446)** | **Controls (n = 276436)** | **Cases (n = 4230)** | **Controls (n = 33541)** | **Cases (n = 611)** | **Controls (n = 27248)** | **Cases (n = 255)** |
| Male (n, %) | 13751 (46.5) | 254 (57.0) | 127823 (46.2) | 2425 (57.3) | 14851 (44.3) | 330 (54.0) | 12746 (46.8) | 128 (50.2) |
| Female (n, %) | 15803 (53.5) | 192 (43.0) | 148611 (53.8) | 1805 (42.7) | 18690 (55.7) | 281 (46.0) | 14502 (53.2) | 127 (49.8) |
| Age (mean, SD) | 56.82 (8.01) | 61.64 (6.10) | 56.84 (7.99) | 61.41 (6.15) | 56.31 (8.05) | 61.00 (6.51) | 52.75 (8.25) | 58.25 (7.97) |
| Age (min-max) | 40-70 | 40-70 | 39-72 | 40-70 | 40-70 | 40-70 | 39-72 | 40-70 |

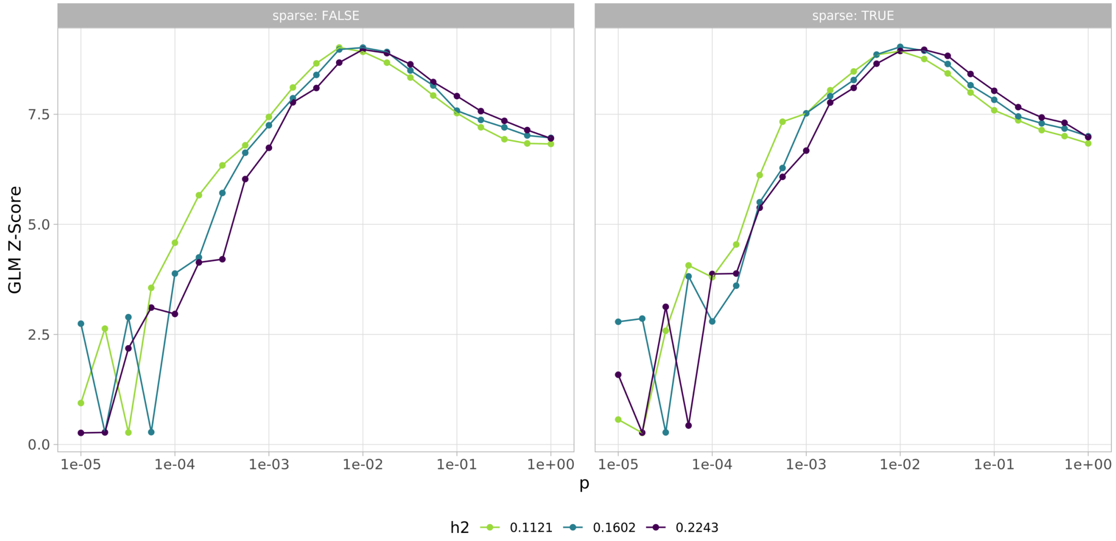

**Figure S2. Z-scores for LDpred2-grid PRS calculated across a grid of tuning parameters: estimated heritability (h2), proportion of causal variants, p, and sparsity (true or false) (after Privé *et al.*)** For the top performing non-sparse grid PRS, the proportion of causal variants was 0.0056, and heritability of 0.1121; for the top-performing sparse model, proportion of causal variants was 0.01, and heritability 0.1602, with sparsity 0.44137.

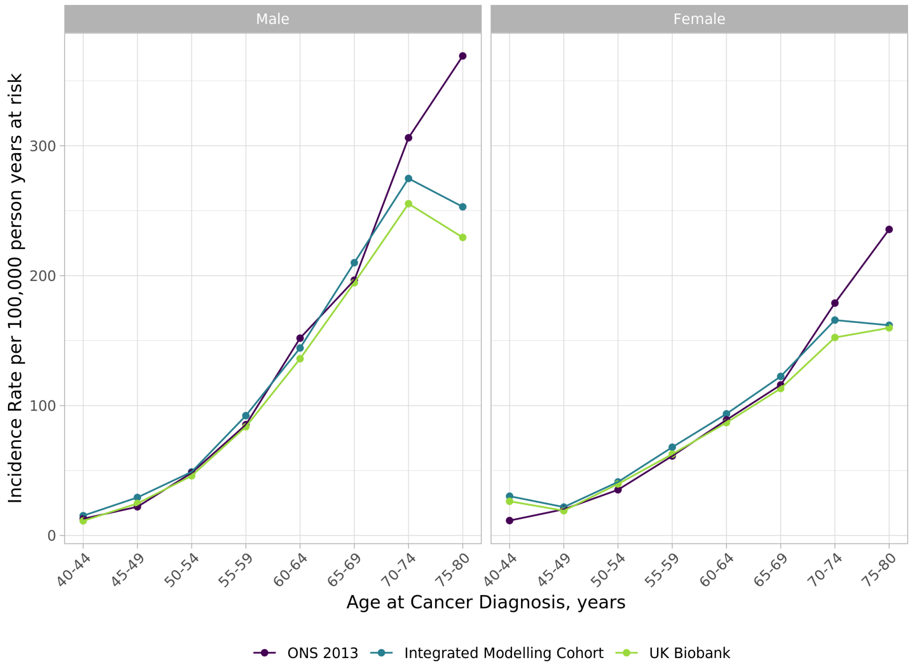

**Figure S3. Age specific CRC rates in men and women in the UK Biobank cohort overall and Integrated Modelling cohorts, compared to Office for National Statistics 2013 Cancer Registry data.**[20] Cases for the whole UK Biobank cohort are from linked cancer registry data; cases for the Integrated Modelling Cohort are from linked cancer registry, death registry, and hospital data.

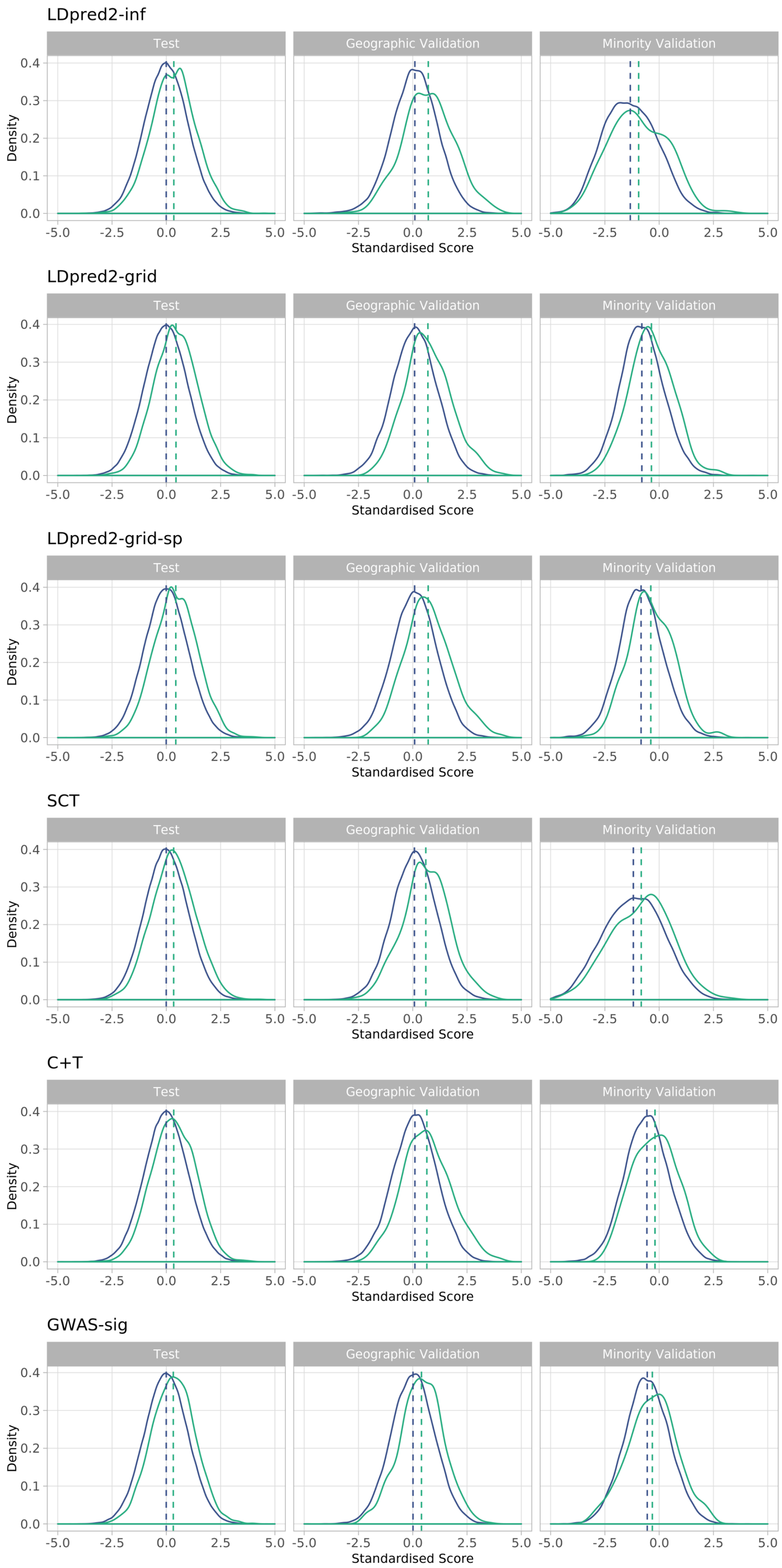

**Figure S4. Distributions of standardised PRS for PRS Test Cohort and Validation Cohorts.** Case distribution is shown in green, controls in blue.

### Interactions between PRS and Age

Evaluation of interaction terms (Table S5) indicated a significant interaction between age and PRS (at $p < 0.01$) for the LDpred-inf model only in logistic regression models, and for LDpred2-grid, LDpred2-grid-sp and C+T Cox models. Plots of marginal effects (shown for logistic regression models in Figure S5) indicated a reduction in effect of PRS with increasing age. Plots for Cox models were similar. Given the weakness of the interaction terms relative to the other predictors based on Wald $\chi^2$, we elected not to include interaction terms in the models.

**Table S5. Wald $\chi^2$ of interaction terms between PRS and age in logistic regression and Cox models**

| | $\chi$2 (p value) | | |
|---|---|---|---|
| | **PRS** | **age** | **PRS * age** |
| **Logistic regression** | | | |
| LDpred2-inf | 529 (<0.001) | 1254 (<0.001) | 8 (0.004) |
| LDpred2-grid | 860 (<0.001) | 1254 (<0.001) | 3 (0.065) |
| LDpred2-grid-sp | 829 (<0.001) | 1254 (<0.001) | 3 (0.068) |
| SCT | 500 (<0.001) | 1254 (<0.001) | 2 (0.136) |
| C+T | 509 (<0.001) | 1252 (<0.001) | 3 (0.064) |
| GWAS-sig | 447 (<0.001) | 1248 (<0.001) | 1 (0.457) |
| **Cox regression** | | | |
| LDpred2-inf | 207 (<0.001) | 575 (<0.001) | 4 (0.038) |
| LDpred2-grid | 428 (<0.001) | 578 (<0.001) | 9 (0.003) |
| LDpred2-grid-sp | 405 (<0.001) | 577 (<0.001) | 8 (0.005) |
| SCT | 222 (<0.001) | 576 (<0.001) | 4 (0.035) |
| C+T | 242 (<0.001) | 576 (<0.001) | 9 (0.003) |
| GWAS-sig | 225 (<0.001) | 574 (<0.001) | 7 (0.011) |

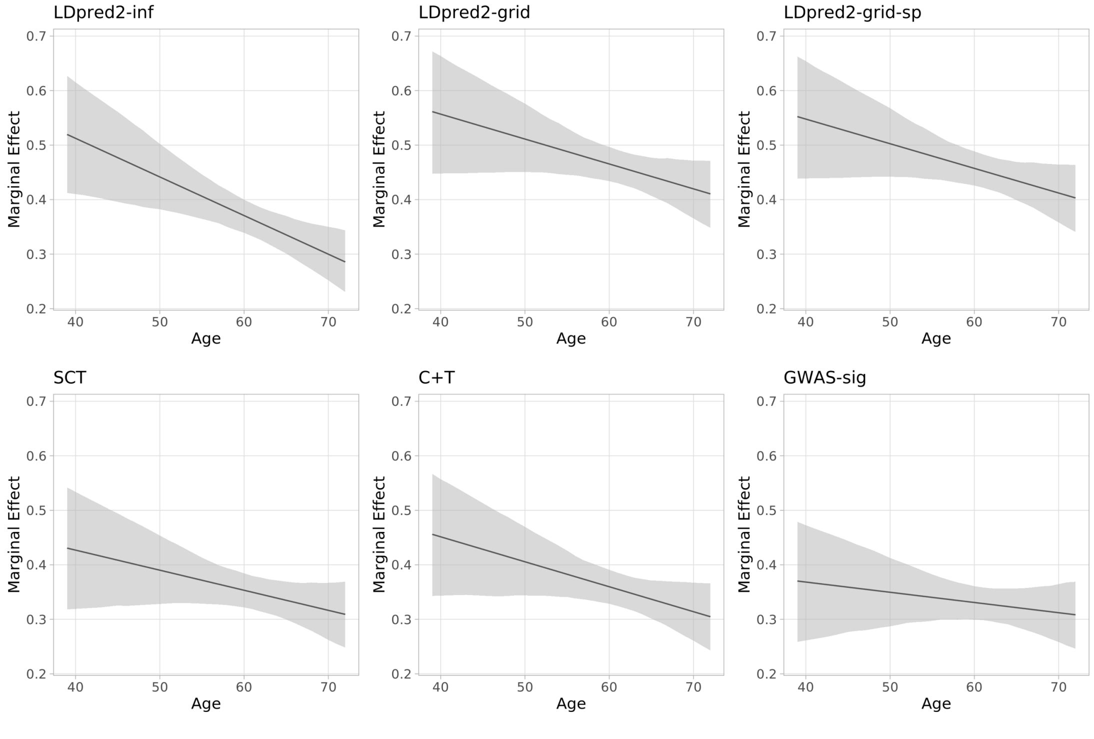

**Figure S5. Marginal effect of standardised PRS in interaction with age in linear regression models.**

**Table S6. Apparent performance of PRS assessed in logistic regression models in the Test Cohort, with and without adjustment for sex and age**

| | LDpred2-inf | LDpred2-grid | LDpred2-grid-sp | SCT | C+T | GWAS-sig |
|---|---|---|---|---|---|---|
| **With sex and age** | | | | | | |
| C | 0.704 (0.697 - 0.712) | 0.717 (0.711 - 0.725) | 0.716 (0.710 - 0.723) | 0.702 (0.695 - 0.711) | 0.704 (0.697 - 0.711) | 0.700 (0.693 - 0.707) |
| Dxy | 0.407 (0.394 - 0.423) | 0.435 (0.422 - 0.451) | 0.432 (0.419 - 0.446) | 0.404 (0.389 - 0.422) | 0.407 (0.394 - 0.423) | 0.400 (0.386 - 0.414) |
| R2 (%) | 5.5 (5.1 - 5.9) | 6.3 (5.9 - 6.8) | 6.2 (5.8 - 6.7) | 5.4 (5.0 - 5.9) | 5.4 (5.1 - 5.9) | 5.3 (4.9 - 5.7) |
| Scaled Brier (%) | 0.87 | 1.05 | 1.03 | 0.86 | 0.85 | 0.83 |
| **Without sex and age** | | | | | | |
| C | 0.597 (0.589 - 0.606) | 0.626 (0.618 - 0.634) | 0.623 (0.614 - 0.632) | 0.594 (0.587 - 0.603) | 0.597 (0.589 - 0.606) | 0.592 (0.584 - 0.601) |
| Dxy | 0.194 (0.178 - 0.212) | 0.251 (0.235 - 0.268) | 0.247 (0.229 - 0.264) | 0.189 (0.175 - 0.206) | 0.193 (0.178 - 0.211) | 0.185 (0.169 - 0.202) |
| R2 (%) | 1.3 (1.1 - 1.5) | 2.1 (1.8 - 2.4) | 2.0 (1.8 - 2.3) | 1.2 (1.0 - 1.5) | 1.3 (1.1 - 1.5) | 1.1 (0.9 - 1.3) |
| Scaled Brier (%) | 0.21 | 0.34 | 0.33 | 0.19 | 0.19 | 0.17 |

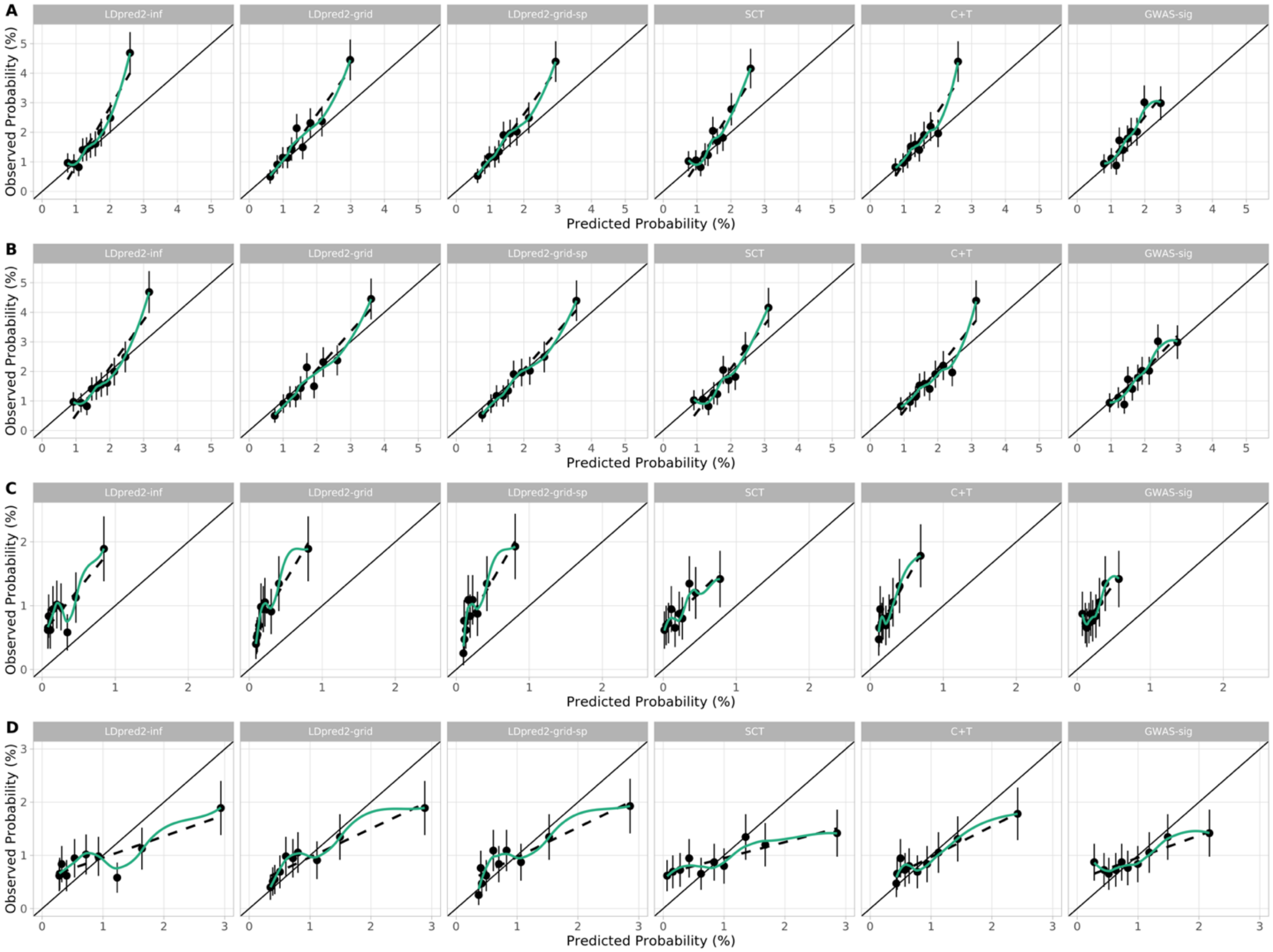

**Figure S6. Calibration plots of PRS models in logistic regression analyses in Validation Cohorts**. Panels show calibration for the Geographic Validation Cohort before (A) and after (B) recalibration, and in the Minority Ethnic Validation Cohort before (C) and after (D) recalibration. Plots show predicted and observed risks by tenths of PRS for each model.

**Table S7. Subgroup analysis of PRS logistic regression model performance by sex and in individuals with a first degree family history of CRC in the Geographic Validation Cohort.**

| | LDpred2-inf | LDpred2-grid | LDpred2-grid-sp | SCT | C+T | GWAS-sig |
|---|---|---|---|---|---|---|
| **Males** | | | | | | |
| C | 0.731 (0.705 - 0.760) | 0.740 (0.716 - 0.767) | 0.741 (0.715 - 0.768) | 0.728 (0.702 - 0.753) | 0.726 (0.702 - 0.755) | 0.716 (0.689 - 0.743) |
| Dxy | 0.463 (0.410 - 0.519) | 0.481 (0.433 - 0.534) | 0.481 (0.431 - 0.536) | 0.455 (0.404 - 0.507) | 0.453 (0.403 - 0.510) | 0.433 (0.378 - 0.486) |
| R2 (%) | 7.6 (6.0 - 9.3) | 8.3 (6.6 - 10.1) | 8.3 (6.6 - 10.1) | 7.2 (5.5 - 8.7) | 7.2 (5.6 - 8.9) | 6.6 (5.0 - 8.2) |
| Slope | 1.216 (1.047 - 1.409) | 1.171 (1.025 - 1.343) | 1.182 (1.034 - 1.357) | 1.178 (1.006 - 1.354) | 1.187 (1.026 - 1.371) | 1.137 (0.968 - 1.320) |
| CITL | 0.186 (0.075 - 0.287) | 0.178 (0.068 - 0.279) | 0.180 (0.070 - 0.281) | 0.174 (0.067 - 0.275) | 0.176 (0.066 - 0.278) | 0.170 (0.061 - 0.273) |
| Scaled Brier (%) | 1.73 | 1.90 | 1.90 | 1.47 | 1.60 | 1.37 |
| **Females** | | | | | | |
| C | 0.709 (0.680 - 0.739) | 0.712 (0.682 - 0.741) | 0.714 (0.684 - 0.743) | 0.694 (0.666 - 0.723) | 0.699 (0.669 - 0.731) | 0.673 (0.643 - 0.703) |
| Dxy | 0.419 (0.360 - 0.477) | 0.423 (0.365 - 0.481) | 0.427 (0.368 - 0.486) | 0.387 (0.331 - 0.447) | 0.397 (0.338 - 0.462) | 0.346 (0.287 - 0.406) |
| R2 (%) | 5.7 (4.0 - 7.4) | 6.0 (4.1 - 7.7) | 6.1 (4.3 - 7.8) | 4.8 (3.2 - 6.6) | 5.2 (3.3 - 6.9) | 3.4 (1.8 - 5.1) |
| Slope | 1.102 (0.919 - 1.292) | 1.035 (0.881 - 1.196) | 1.055 (0.896 - 1.214) | 1.002 (0.839 - 1.185) | 1.041 (0.863 - 1.236) | 0.862 (0.700 - 1.036) |
| CITL | 0.230 (0.111 - 0.352) | 0.221 (0.100 - 0.343) | 0.223 (0.101 - 0.344) | 0.217 (0.098 - 0.339) | 0.218 (0.098 - 0.340) | 0.215 (0.097 - 0.342) |
| Scaled Brier (%) | 1.04 | 1.20 | 1.22 | 0.90 | 0.98 | 0.59 |
| **First degree family history** | | | | | | |
| C | 0.697 (0.637 - 0.748) | 0.701 (0.642 - 0.754) | 0.706 (0.647 - 0.758) | 0.685 (0.625 - 0.738) | 0.703 (0.646 - 0.752) | 0.668 (0.608 - 0.721) |
| Dxy | 0.394 (0.275 - 0.496) | 0.402 (0.283 - 0.509) | 0.412 (0.293 - 0.515) | 0.369 (0.251 - 0.475) | 0.406 (0.292 - 0.504) | 0.335 (0.217 - 0.443) |
| R2 (%) | 2.4 (-2.2 - 6.0) | 3.4 (-1.1 - 7.4) | 3.6 (-0.9 - 7.5) | 1.8 (-2.8 - 5.8) | 2.9 (-1.7 - 6.6) | 0.5 (-4.1 - 4.6) |
| Slope | 1.021 (0.714 - 1.322) | 0.971 (0.683 - 1.259) | 0.997 (0.714 - 1.286) | 0.948 (0.633 - 1.246) | 1.052 (0.745 - 1.363) | 0.838 (0.518 - 1.145) |
| CITL | 0.658 (0.462 - 0.827) | 0.607 (0.409 - 0.771) | 0.613 (0.414 - 0.776) | 0.643 (0.451 - 0.812) | 0.644 (0.453 - 0.810) | 0.633 (0.443 - 0.809) |
| Scaled Brier (%) | 1.07 | 1.45 | 1.45 | 1.03 | 1.14 | 0.77 |

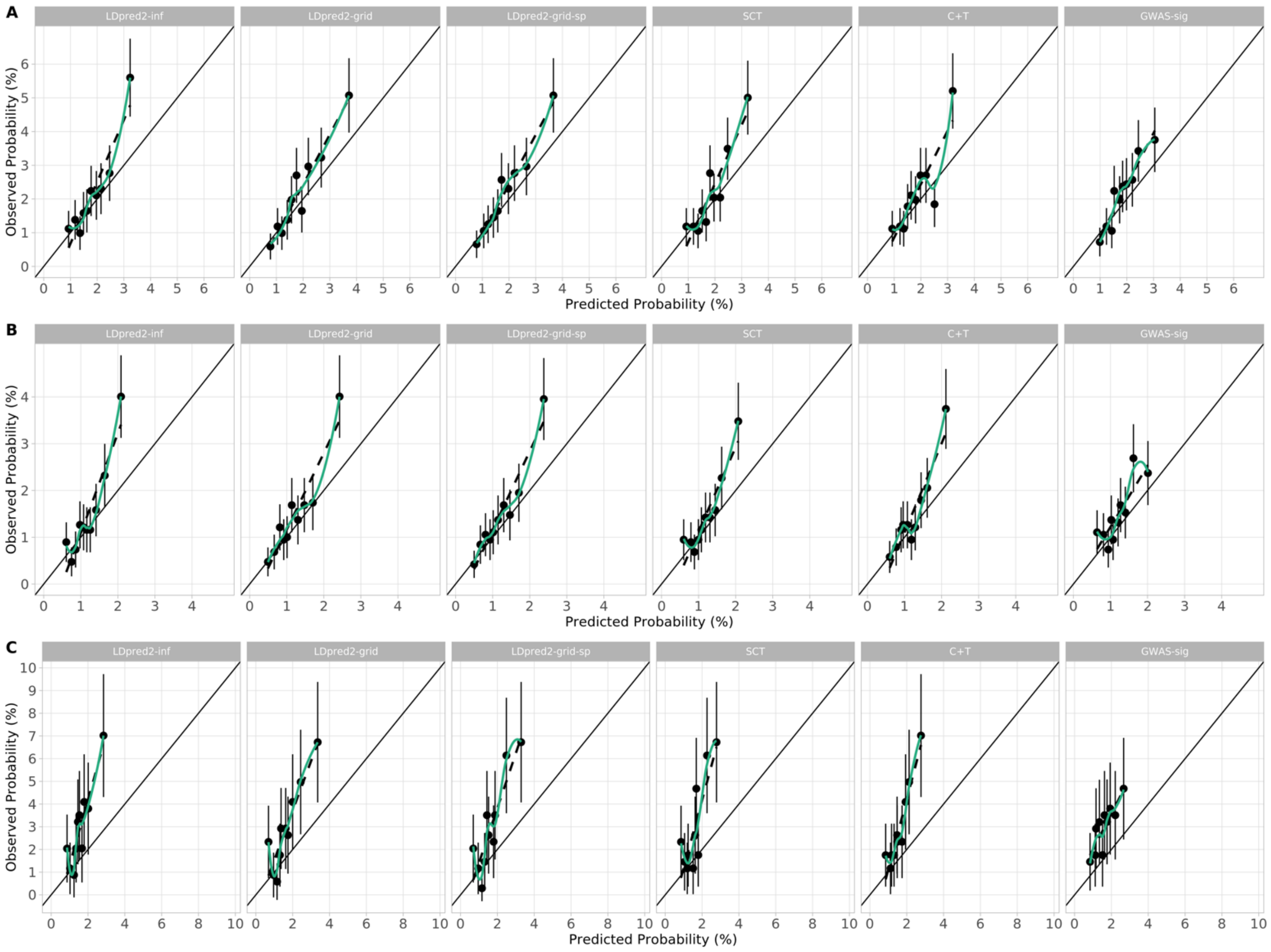

**Figure S7. Calibration of PRS in logistic regression models in subgroup analysis in the Geographic Validation Cohort.** Plots show predicted and observed risks by tenths of PRS for each model in males (A), females (B), and those with a first degree family history of CRC (C).

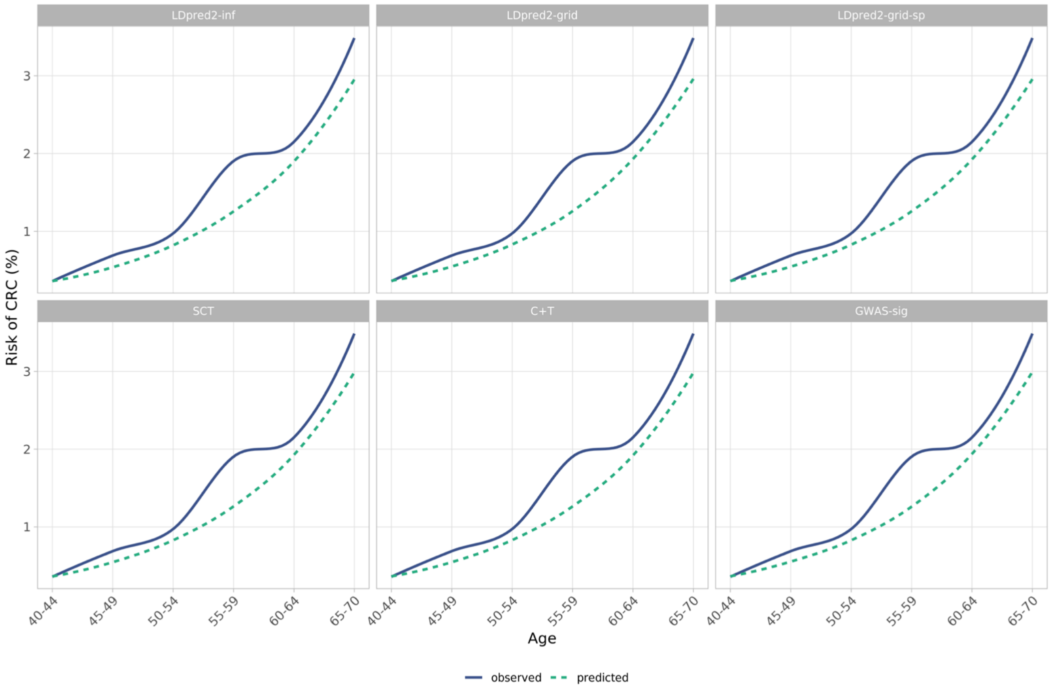

**Figure S8. Observed and predicted probabilities of CRC for PRS logistic regression models across 5 year age bands in the Geographic Validation Cohort.**

**Table S8. Apparent, internally and externally validated polygenic risk score (PRS) performance in Cox's proportional hazards models (adjusting for age, sex, array and first 4 principal components).** Values are performance indices plus 95% confidence intervals are provided for each cohort. PRS HR per SD – adjusted hazard ratio of PRS in model per standard deviation of the PRS; C – Harrell's C index; Dxy – Somers' Dxy rank correlation; D – Royston's D statistic; R2D – Royston and Sauerbrei's $R_D^2$ (explained variation); Slope – Calibration Slope. Pairwise comparison of performance metrics in validation cohorts (paired t-tests with Bonferroni correction) were all significantly different P<0.001 except comparisons marked ^/* where P>0.20.

| | LDpred2-inf | LDpred2-grid | LDpred2-grid-sp | SCT | C+T | GWAS-sig | Reference |
|---|---|---|---|---|---|---|---|
| **Apparent performance** | | | | | | | |
| PRS HR per SD | 1.368 (1.310 - 1.428) | 1.563 (1.498 - 1.631) | 1.545 (1.480 - 1.612) | 1.378 (1.321 - 1.438) | 1.397 (1.338 - 1.459) | 1.377 (1.320 - 1.436) | NA |
| C | 0.696 (0.685 - 0.707) | 0.714 (0.704 - 0.726) | 0.712 (0.702 - 0.723) | 0.695 (0.685 - 0.706) | 0.698 (0.689 - 0.709) | 0.695 (0.685 - 0.706) | 0.675 (0.665 - 0.687) |
| Dxy | 0.391 (0.370 - 0.414) | 0.427 (0.409 - 0.451) | 0.424 (0.403 - 0.447) | 0.391 (0.370 - 0.412) | 0.396 (0.378 - 0.417) | 0.390 (0.370 - 0.412) | 0.350 (0.331 - 0.373) |
| D | 1.085 (1.027 - 1.150) | 1.201 (1.143 - 1.268) | 1.190 (1.132 - 1.255) | 1.096 (1.034 - 1.163) | 1.099 (1.043 - 1.162) | 1.094 (1.031 - 1.162) | 0.961 (0.902 - 1.021) |
| R2D (%) | 22.0 (20.1 - 24.0) | 25.6 (23.8 - 27.8) | 25.3 (23.4 - 27.3) | 22.3 (20.3 - 24.4) | 22.4 (20.6 - 24.4) | 22.2 (20.2 - 24.4) | 18.1 (16.3 - 19.9) |
| Scaled Brier (% | 0.45 | 0.56 | 0.55 | 0.49 | 0.47 | 0.50 | 0.39 |
| **Internal validation** | | | | | | | |
| C | 0.694 | 0.713 | 0.711 | 0.694 | 0.697 | 0.694 | 0.674 |
| Dxy | 0.389 | 0.425 | 0.422 | 0.389 | 0.393 | 0.387 | 0.347 |
| D | 1.078 | 1.194 | 1.183 | 1.089 | 1.091 | 1.088 | 0.954 |
| R2D | 21.7 | 25.4 | 25.1 | 22.1 | 22.1 | 22.0 | 17.8 |
| Slope | 0.992 | 0.994 | 0.995 | 0.994 | 0.992 | 0.992 | 0.992 |
| Scaled Brier (% | 0.44 | 0.55 | 0.54 | 0.47 | 0.46 | 0.49 | 0.38 |
| **Geographic Validation** | | | | | | | |
| C | 0.715 (0.686 - 0.743) | 0.724 (0.696 - 0.751) | 0.725 (0.696 - 0.752) | 0.713 (0.686 - 0.740) | 0.707 (0.681 - 0.734) | 0.701 (0.675 - 0.729) | 0.673 (0.644 - 0.702) |
| Dxy | 0.430 (0.372 - 0.485) | 0.448 (0.391 - 0.501) | 0.450 (0.393 - 0.504) | 0.426 (0.372 - 0.480) | 0.415 (0.361 - 0.468) | 0.402 (0.350 - 0.458) | 0.345 (0.288 - 0.404) |
| D | 1.243 (1.075 - 1.406) | 1.285 (1.124 - 1.448) | 1.293 (1.130 - 1.460) | 1.184 (1.029 - 1.346)^ | 1.182 (1.023 - 1.348)^ | 1.145 (0.992 - 1.319) | 0.945 (0.790 - 1.113) |
| R2D | 27.0 (21.7 - 32.1) | 28.3 (23.2 - 33.3) | 28.5 (23.4 - 33.7) | 25.1 (20.2 - 30.2)^ | 25.1 (20.0 - 30.3)^ | 23.8 (19.0 - 29.4) | 17.6 (13.0 - 22.9) |
| Slope | 1.123 (0.950 - 1.291) | 1.058 (0.911 - 1.204)^ | 1.073 (0.925 - 1.220)* | 1.070 (0.919 - 1.234)* | 1.054 (0.897 - 1.223)^ | 1.023 (0.869 - 1.204) | 0.947 (0.774 - 1.142) |
| Scaled Brier (% | 0.75 | 0.76 | 0.78 | 0.63 | 0.61 | 0.59 | 0.37 |
| **Minority Ethnic Validation** | | | | | | | |
| C | 0.647 (0.593 - 0.700)^ | 0.666 (0.610 - 0.720) | 0.664 (0.609 - 0.718) | 0.650 (0.596 - 0.705) | 0.658 (0.606 - 0.710) | 0.659 (0.605 - 0.715) | 0.647 (0.595 - 0.702)^ |
| Dxy | 0.293 (0.185 - 0.399)^ | 0.331 (0.221 - 0.440) | 0.329 (0.219 - 0.437) | 0.300 (0.192 - 0.410) | 0.316 (0.212 - 0.420) | 0.319 (0.210 - 0.430) | 0.293 (0.189 - 0.403)^ |
| D | 0.931 (0.650 - 1.273) | 1.033 (0.736 - 1.374) | 1.030 (0.734 - 1.363) | 0.940 (0.640 - 1.281) | 0.981 (0.682 - 1.320) | 0.995 (0.693 - 1.335) | 0.889 (0.610 - 1.229) |
| R2D | 17.2 (9.2 - 27.9) | 20.3 (11.5 - 31.1) | 20.2 (11.4 - 30.7) | 17.4 (8.9 - 28.1) | 18.7 (10.0 - 29.4)^ | 19.1 (10.3 - 29.9)^ | 15.9 (8.1 - 26.5) |
| Slope | 0.262 (0.161 - 0.397) | 0.314 (0.205 - 0.452) | 0.318 (0.207 - 0.455) | 0.252 (0.154 - 0.384)^ | 0.297 (0.188 - 0.442) | 0.251 (0.151 - 0.389)^ | 0.232 (0.136 - 0.366) |
| Scaled Brier (% | 0.16 | 0.26 | 0.26 | 0.16 | 0.21 | 0.19 | 0.14 |

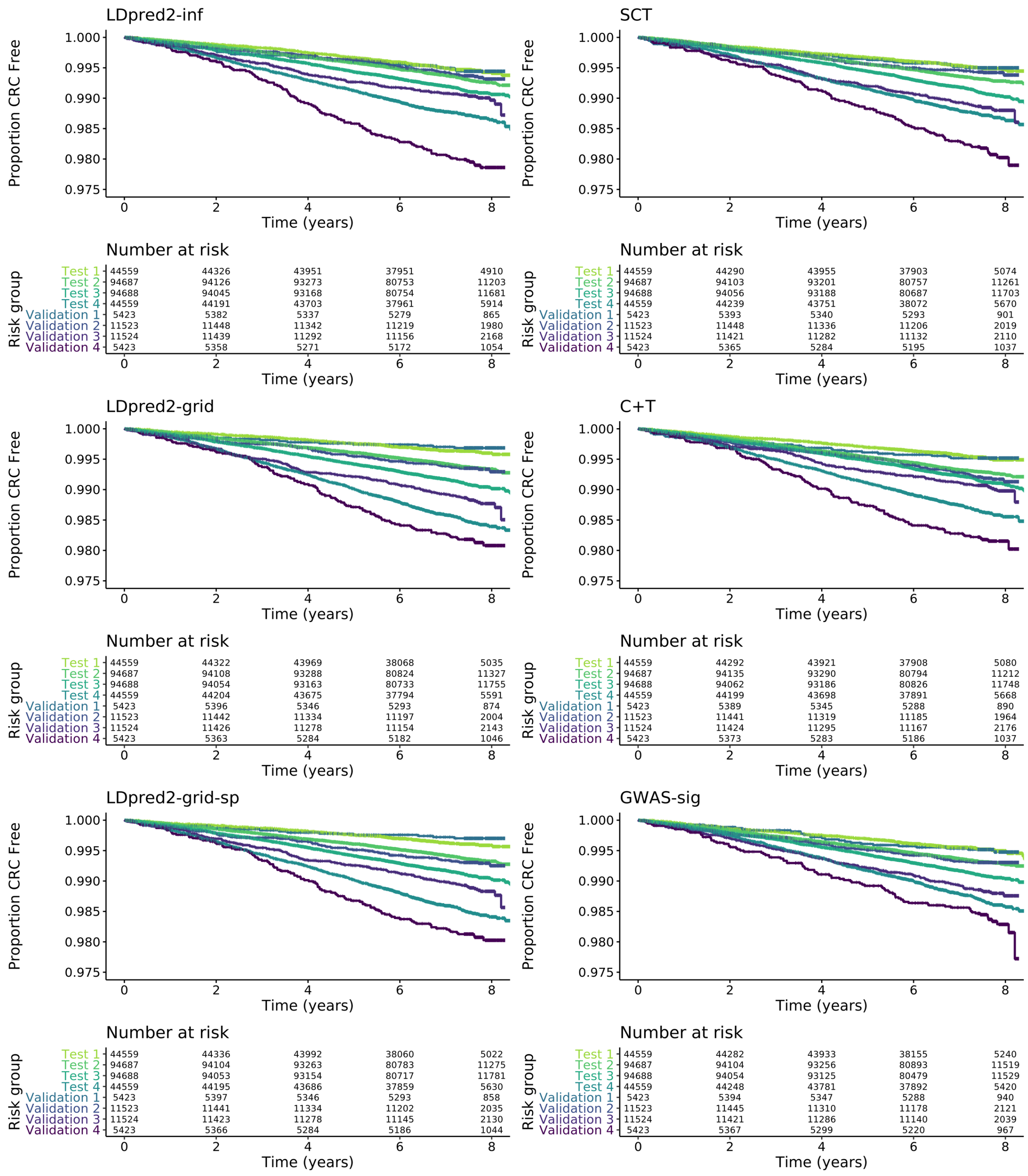

**Figure S9. Kaplan-Meier curves across four risk groups (group 4 being highest risk) for PRS in the Geographic Validation Cohort compared with the Test Cohort**

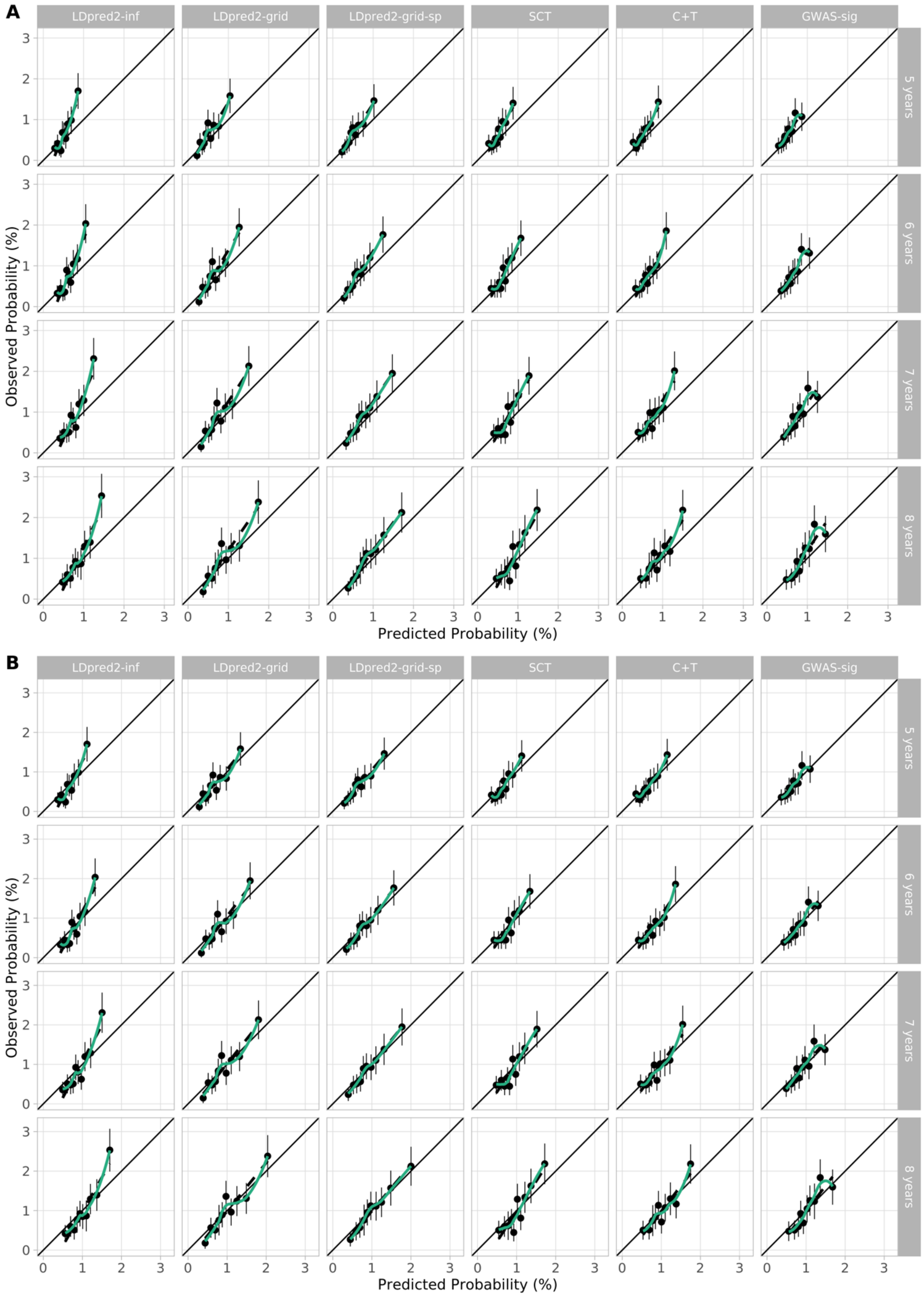

**Figure S10. Calibration plots of PRS models in Cox models in the Geographic Validation Cohort**. Plots show predicted and observed risks by tenths of PRS before (A) and after (B) recalibration for each model.

**Table S9. Subgroup analysis of PRS Cox model performance by sex in the Geographic Validation Cohort.** We did not assess performance specifically in those with a first degree family history, as there were too few incident cases in this group.

| | LDpred2-inf | LDpred2-grid | LDpred2-grid-sp | SCT | C+T | GWAS-sig |
|---|---|---|---|---|---|---|
| **Males** | | | | | | |
| C | 0.709 (0.675 - 0.747) | 0.724 (0.691 - 0.761) | 0.723 (0.691 - 0.760) | 0.711 (0.677 - 0.745) | 0.704 (0.668 - 0.740) | 0.707 (0.675 - 0.740) |
| Dxy | 0.419 (0.349 - 0.493) | 0.448 (0.382 - 0.522) | 0.446 (0.382 - 0.520) | 0.422 (0.354 - 0.489) | 0.408 (0.337 - 0.481) | 0.414 (0.350 - 0.481) |
| D | 1.197 (0.989 - 1.430) | 1.272 (1.072 - 1.486) | 1.271 (1.063 - 1.499) | 1.149 (0.963 - 1.359) | 1.156 (0.938 - 1.383) | 1.185 (0.991 - 1.394) |
| R2D (%) | 25.5 (18.9 - 32.8) | 27.9 (21.5 - 34.5) | 27.8 (21.2 - 34.9) | 24.0 (18.1 - 30.6) | 24.1 (17.3 - 31.3) | 25.1 (19.0 - 31.7) |
| Slope | 1.172 (0.954 - 1.431) | 1.120 (0.942 - 1.327) | 1.128 (0.944 - 1.350) | 1.139 (0.947 - 1.370) | 1.117 (0.896 - 1.365) | 1.157 (0.950 - 1.390) |
| Scaled Brier (%) | 0.82 | 0.84 | 0.85 | 0.67 | 0.67 | 0.72 |
| **Females** | | | | | | |
| C | 0.707 (0.670 - 0.745) | 0.711 (0.671 - 0.746) | 0.713 (0.673 - 0.749) | 0.700 (0.657 - 0.738) | 0.696 (0.655 - 0.731) | 0.680 (0.638 - 0.720) |
| Dxy | 0.414 (0.340 - 0.490) | 0.421 (0.342 - 0.492) | 0.427 (0.345 - 0.498) | 0.399 (0.313 - 0.476) | 0.393 (0.309 - 0.461) | 0.360 (0.276 - 0.439) |
| D | 1.244 (0.994 - 1.492) | 1.227 (0.985 - 1.459) | 1.250 (1.005 - 1.485) | 1.142 (0.882 - 1.398) | 1.133 (0.887 - 1.377) | 1.004 (0.769 - 1.245) |
| R2D (%) | 27.0 (19.1 - 34.7) | 26.432 (18.8 - 33.7) | 27.2 (19.4 - 34.5) | 23.8 (15.7 - 31.8) | 23.5 (15.8 - 31.1) | 19.4 (12.4 - 27.0) |
| Slope | 1.171 (0.912 - 1.460) | 1.053 (0.827 - 1.278) | 1.080 (0.847 - 1.304) | 1.076 (0.822 - 1.354) | 1.058 (0.802 - 1.312) | 0.944 (0.708 - 1.203) |
| Scaled Brier (%) | 0.55 | 0.52 | 0.56 | 0.44 | 0.41 | 0.30 |

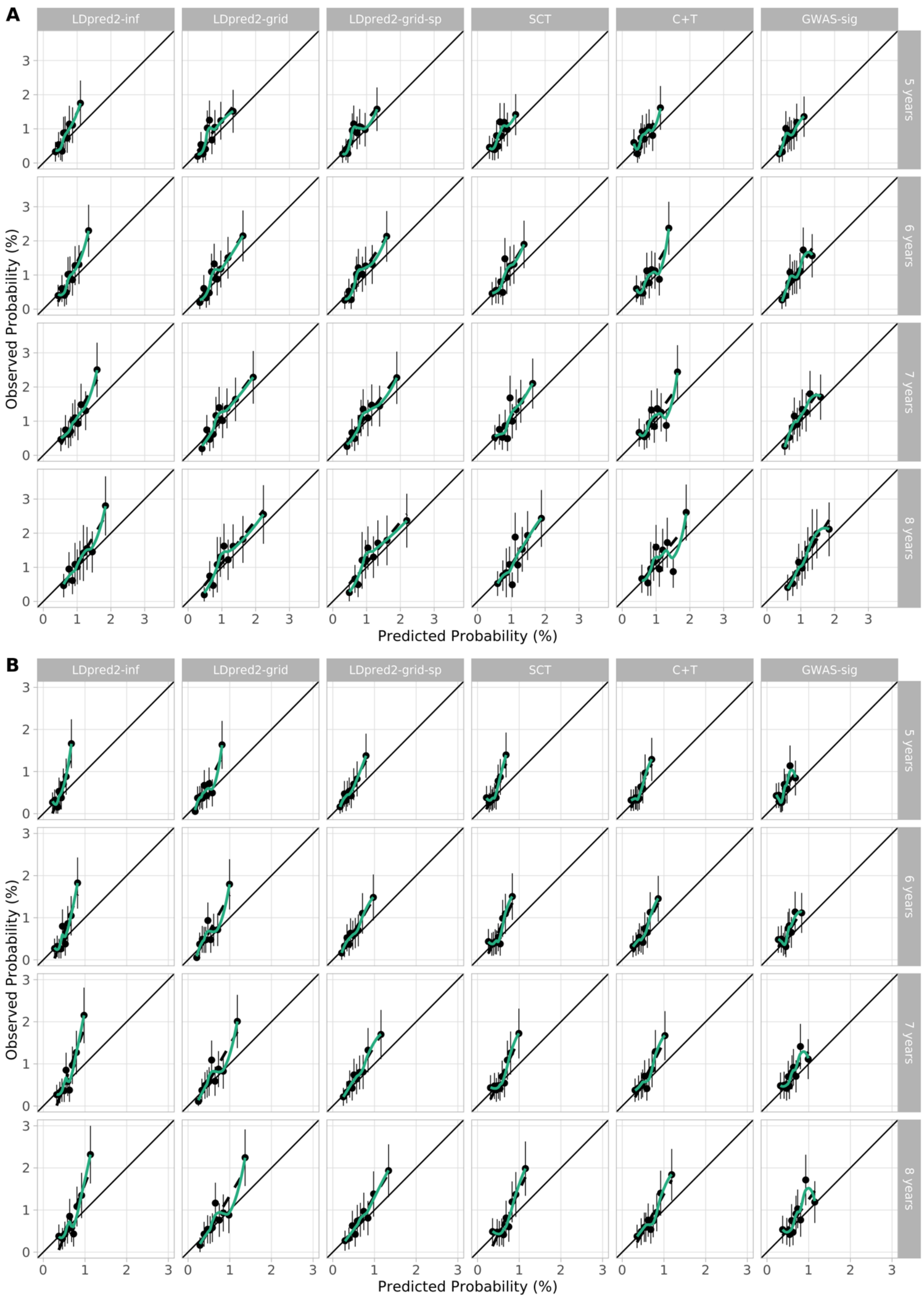

**Figure S11. Calibration of PRS in Cox models by sex in the Geographic Validation Cohort.** Plots show predicted and observed risks by tenths of PRS for each model in males (A) and females (B).

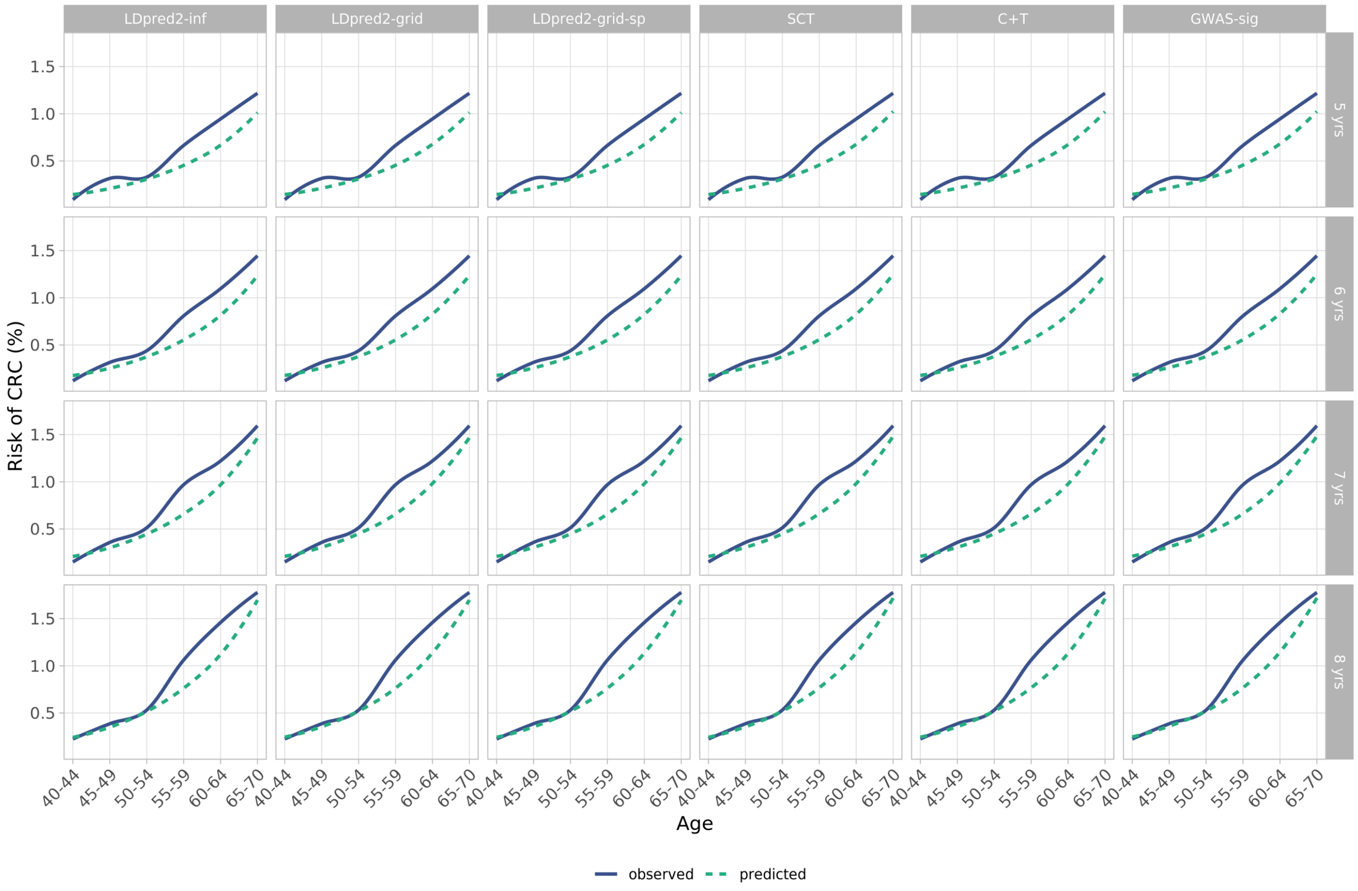

**Figure S12. Observed and predicted probabilities of CRC for PRS Cox models across 5 year age bands in the Geographic Validation Cohort**

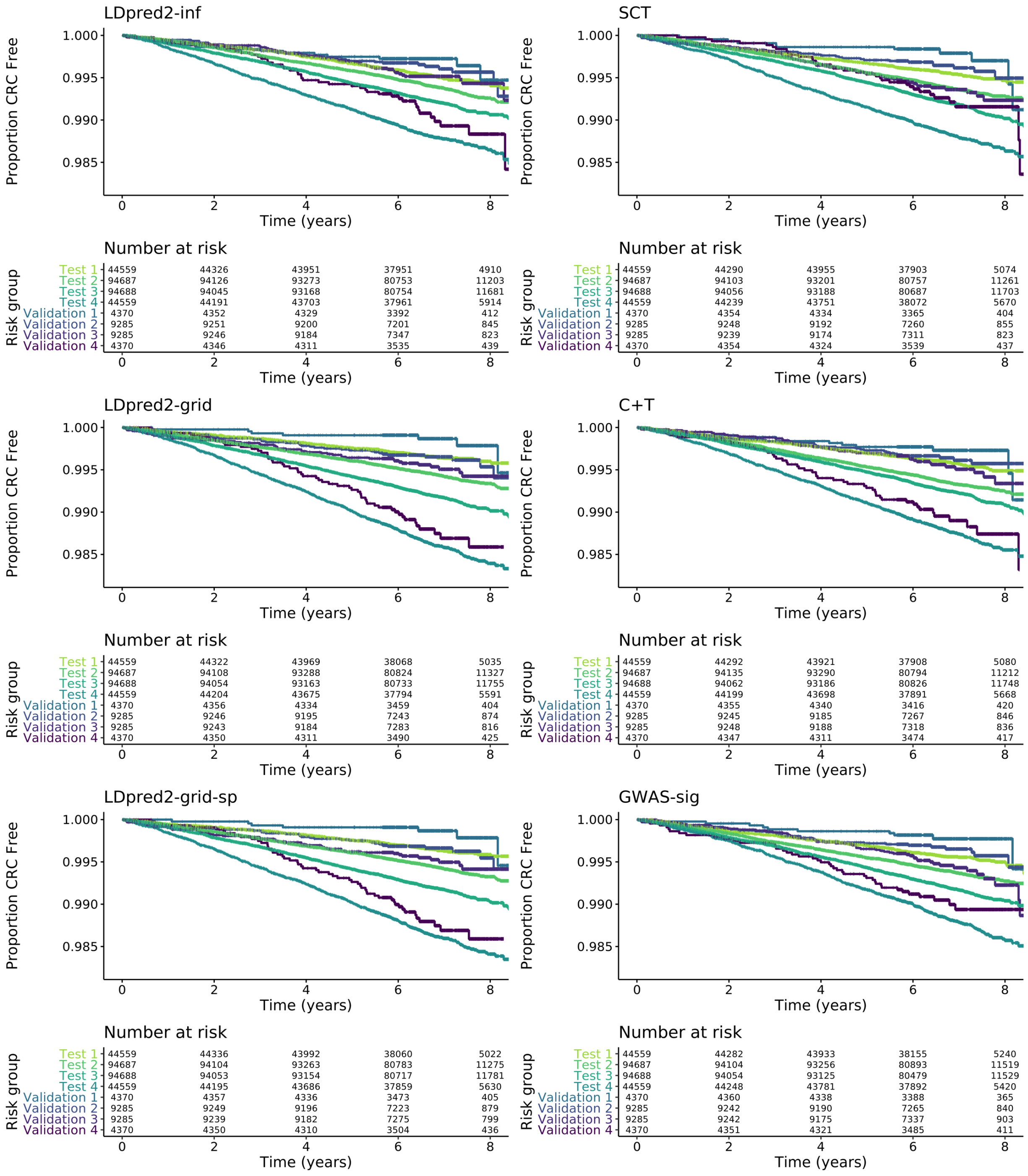

**Figure S13. Kaplan-Meier curves across four risk groups (group 4 being highest risk) for PRS in the Minority Ethnic Validation Cohort compared with the Test Cohort**

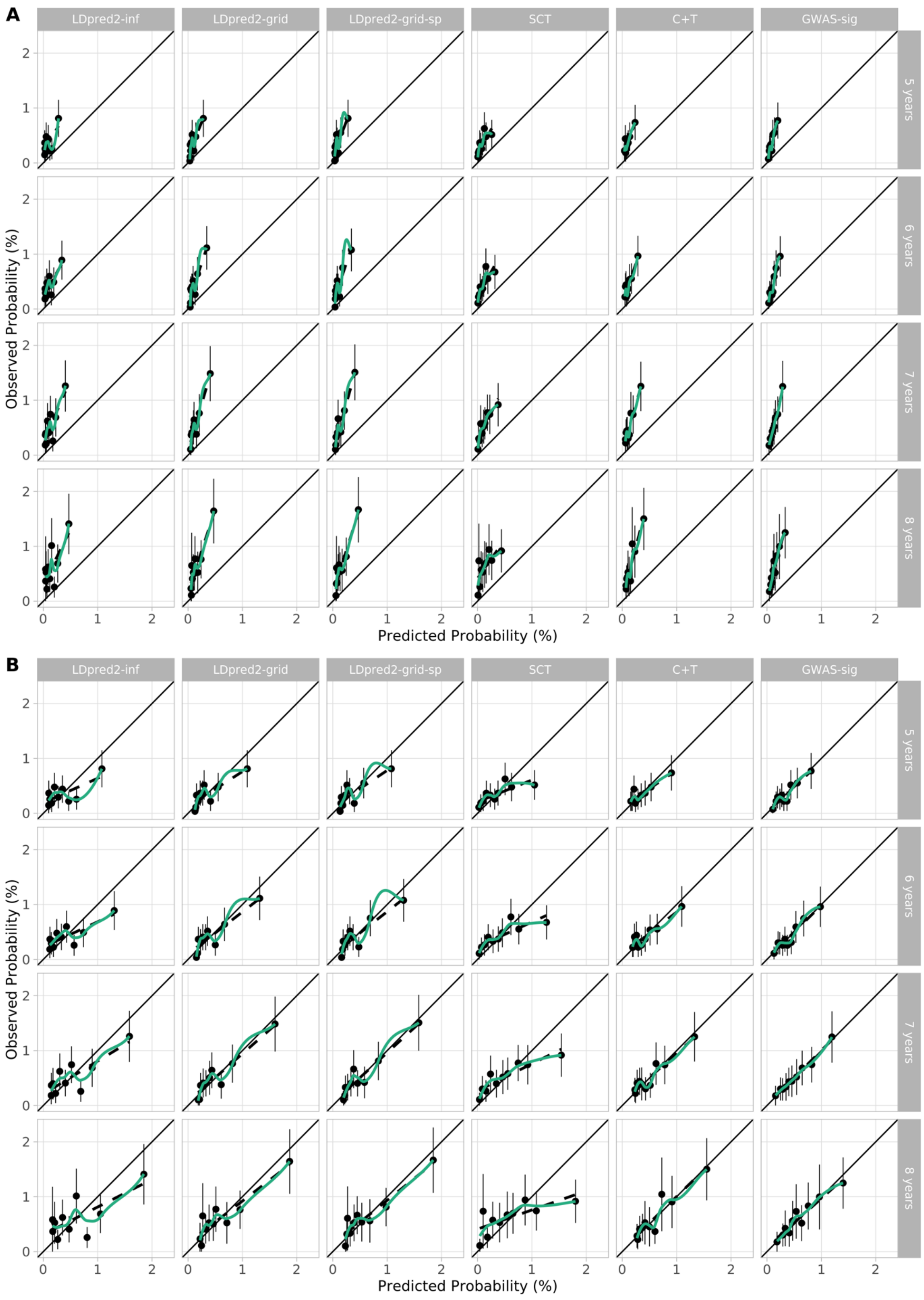

**Figure S14. Calibration plots of PRS models in Cox model in the Minority Ethnic Validation Cohort.** Plots show predicted and observed risks by tenths of PRS cohort before (A) and after (B) recalibration for each model.

**Table S10. Characteristics of the UKB Integrated Modelling Cohort used for QCancer-10 validation, compared with the QCancer-10 derivation cohort.** Values are numbers (%) unless otherwise indicated. CRC – colorectal cancer, NA – not applicable.

| | Male UKB cohort (n = 196091) | Male QCancer-10 derivation (n =2 447 866) | Female UKB cohort (n = 238946) | Female QCancer-10 Derivation (n =2 495 899) |
|---|---|---|---|---|
| Age (years), mean (SD) | 56.7 (8.2) | 44.3 (14.8) | 56.3 (8.0) | 44.9 (15.9) |
| Ethnicity | | | | |
| White/not recorded | 185813 (94.8) | 2 231 641 (91.2) | 224316 (94.6) | 2 271 520 (91.0) |
| Indian | 2510 (1.3) | 42 771 (1.7) | 2601 (1.1) | 37 773 (1.5) |
| Pakistani | 903 (0.5) | 17 169 (0.7) | 616 (0.3) | 16 893 (0.7) |
| Bangladeshi | 132 (0.1) | 17 169 (0.7) | 61 (0.0) | 13 170 (0.5) |
| Other Asian | 841 (0.4) | 24 494 (1.0) | 748 (0.3) | 27 750 (1.1) |
| Caribbean | 1397 (0.7) | 37 003 (1.5) | 1412 (0.6) | 40 742 (1.6) |
| Black African | 1363 (0.7) | 18 553 (0.8) | 2498 (1.0) | 23 920 (1.0) |
| Chinese | 516 (0.3) | 12 493 (0.5) | 865 (0.4) | 17 702 (0.7) |
| Other | 2616 (1.3) | 41 738 (1.7) | 3980 (1.7) | 46 429 (1.9) |
| Townsend deprivation index, mean (SD) | -1.3 (3.1) | 0.3 (3.6) | -1.4 (3.0) | 0.2 (3.6) |
| BMI (kg/m2), mean (SD) | 27.8 (4.2) | 26.3 (4.2) | 27.0 (5.2) | 25.7 (5.0) |
| Smoking status | | | | |
| Non-smoker | 97088 (49.5) | 1 081 822 (44.2) | 142569 (59.8) | 1 433 446 (57.4) |
| Ex-smoker | 75100 (38.3) | 448 480 (18.3) | 74934 (31.4) | 392 870 (15.7) |
| Light smoker | 9361 (4.8) | 351 559 (14.4) | 8885 (3.7) | 284 482 (11.4) |
| Moderate smoker | 5816 (3.0) | 167 089 (6.8) | 7235 (3.0) | 152 115 (6.1) |
| Heavy smoker | 8726 (4.4) | 139 985 (5.7) | 4873 (2.0) | 86 114 (3.5) |
| Alcohol intake | | | | |
| Non-drinker | 11985 (6.1) | 433 515 (17.7) | 22415 (9.4) | 753 150 (30.2) |
| Trivial drinker | 41810 (21.3) | 585 589 (23.9) | 96085 (40.3) | 849 734 (34.0) |
| Light drinker | 57817 (29.5) | 358 713 (14.7) | 76942 (32.3) | 295 009 (11.8) |
| Moderate drinker | 60694 (31.0) | 486 003 (19.9) | 37830 (15.9) | 176 644 (7.1) |
| Heavy drinker | 14960 (7.6) | 41 223 (1.7) | 3797 (1.6) | 5332 (0.2) |
| Very heavy drinker | 8825 (4.5) | 18 473 (0.8) | 1427 (0.6) | 3743 (0.1) |
| Medical history | | | | |
| Ulcerative colitis | 1053 (0.5) | 8956 (0.4) | 1211 (0.5) | 8983 (0.4) |
| Colorectal polyps | 616 (0.3) | 3146 (0.1) | 612 (0.3) | 2447 (0.1) |
| Diabetes | 12893 (6.6) | 68 727 (2.8) | 7885 (3.3) | 53 070 (2.1) |
| Breast cancer | NA | NA | 9448 (4.0) | 25 108 (1.0) |
| Uterine cancer | NA | NA | 1030 (0.4) | 1987 (0.1) |
| Ovarian cancer | NA | NA | 724 (0.3) | 2242 (0.1) |
| Cervical cancer | NA | NA | 1711 (0.7) | 3582 (0.1) |
| Lung cancer | 125 (0.1) | 1488 (0.1) | NA | NA |
| Blood cancers | 1146 (0.6) | 5953 (0.2) | NA | NA |
| Oral cancer | 483 (0.2) | 964 (0.0) | NA | NA |
| Family history of CRC | 19505 (9.9) | 29 877 (1.2) | 22252 (9.3) | 43 741 (1.8) |

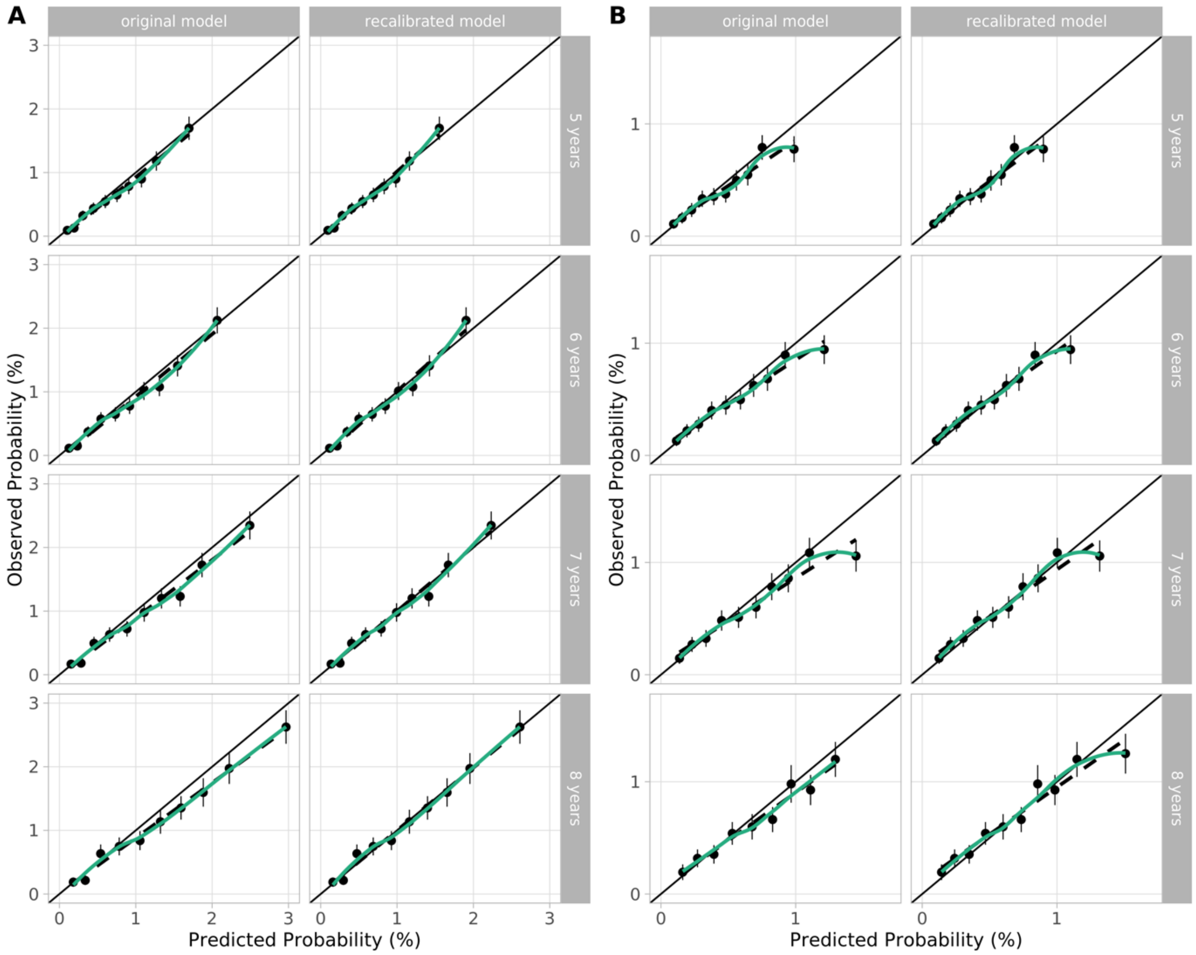

**Figure S15. Calibration of QCancer-10 over 5-8 years of follow-up.** Plots show predicted and observed risks by tenths of predicted risk in males (A) and females (B) before and after recalibration.

**Table S11. Expected/observed ratio of risk over 5-8 years of follow-up for male and female for QCancer-10+LDP, QCancer-10+GWS and QCancer-10 models in subgroups analyses.**

| | Years of follow-up | QCancer-10+LDP | QCancer-10+GWS | QCancer-10 |
|---|---|---|---|---|
| **Family history of CRC** | | | | |
| Male | 5 | 1.06 | 1.05 | 1.02 |
| | 6 | 1.04 | 1.02 | 0.99 |
| | 7 | 1.08 | 1.06 | 1.04 |
| | 8 | 0.97 | 0.95 | 0.93 |
| Female | 5 | 1.28 | 1.25 | 1.31 |
| | 6 | 1.22 | 1.19 | 1.25 |
| | 7 | 1.26 | 1.23 | 1.30 |
| | 8 | 1.19 | 1.16 | 1.23 |
| **Minority ethnicity** | | | | |
| Male | 5 | 0.50 | 0.60 | 0.73 |
| | 6 | 0.57 | 0.68 | 0.82 |
| | 7 | 0.60 | 0.71 | 0.86 |
| | 8 | 0.70 | 0.83 | 1.01 |
| Female | 5 | 0.65 | 0.73 | 0.74 |
| | 6 | 0.58 | 0.65 | 0.65 |
| | 7 | 0.54 | 0.61 | 0.62 |
| | 8 | 0.49 | 0.55 | 0.56 |

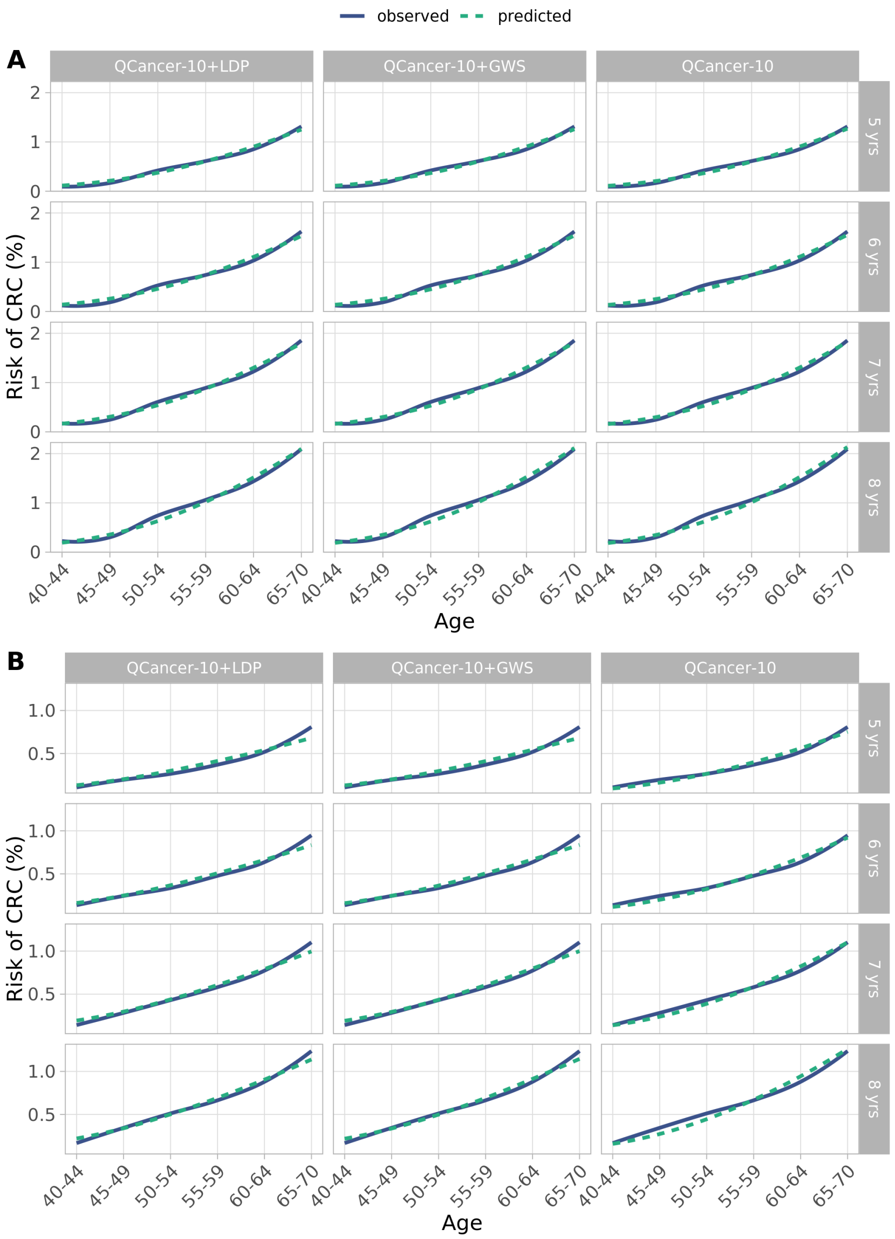

**Figure S16. Observed and predicted probabilities of CRC by age for male and female for QCancer-10+LDP, QCancer-10+GWS and QCancer-10 models.**

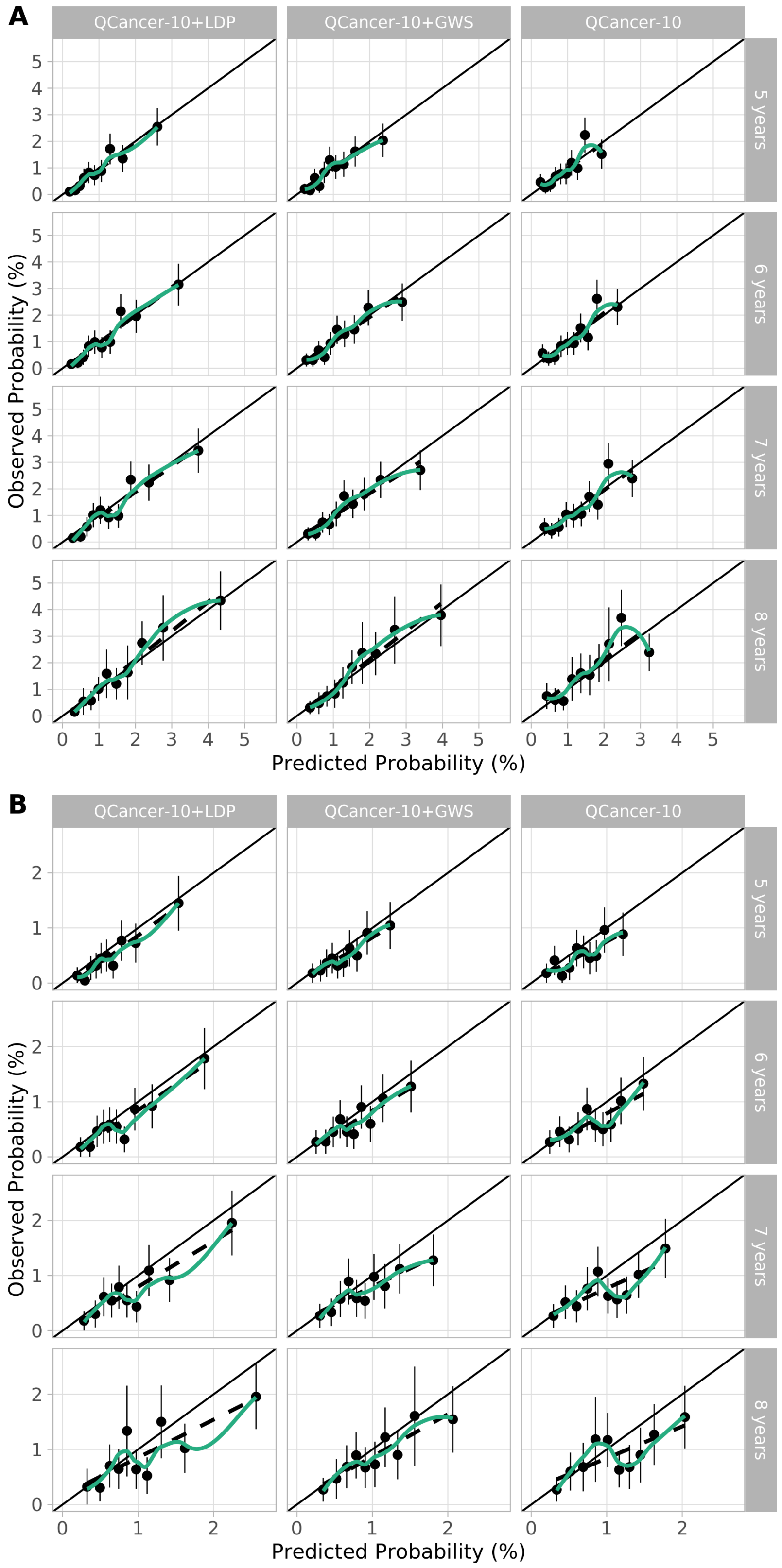

**Figure S17. Calibration plots for individuals with a first-degree family history of CRC in QCancer-10+PRS and QCancer-10 models.** Plots show predicted and observed risks by tenths of predicted risk in males (A) and females (B)

**QCancer-10+PRS model specification**

We confirmed QCancer-10 risk score and PRS fulfilled proportional hazards assumptions. Evaluation of multiple fractional polynomials (MFP) for modelling of these predictors resulted in use of MFP terms for the PRS in the female QCancer-10+LDP model (see Model specification below). Evaluation of interaction terms (Table S11) indicated a significant interaction (at $p < 0.01$) for the male QCancer-10+GWS model only. Plots of marginal effects (Figure S18) indicated a reduction in effect of QCancer-10 with increasing PRS score. Given the weakness of the interaction terms relative to the other predictors based on Wald $\chi^2$, we elected not to include interaction terms in the models.

**Table S12. Interaction terms in QCancer-10+LDP and QCancer-10+GWS models.** Evaluated using MFP terms for female QCancer-10+LDP model

| | QCancer-10+LDP | QCancer-10+GWS |
|---|---|---|
| **Male** | | |
| QCancer-10 LP | 635.85 (<0.001) | 652.88 (<0.001) |
| PRS | 390.82 (<0.001) | 264.36 (<0.001) |
| Interaction term | 6.29 (0.012) | 8.50 (0.004) |
| **Female** | | |
| QCancer-10 LP | 293.43 (<0.001) | 302.90 (<0.001) |
| PRS | 297.96 (<0.001) | 135.83 (<0.001) |
| Interaction term | 0.15 (0.699) | 1.53 (0.216) |

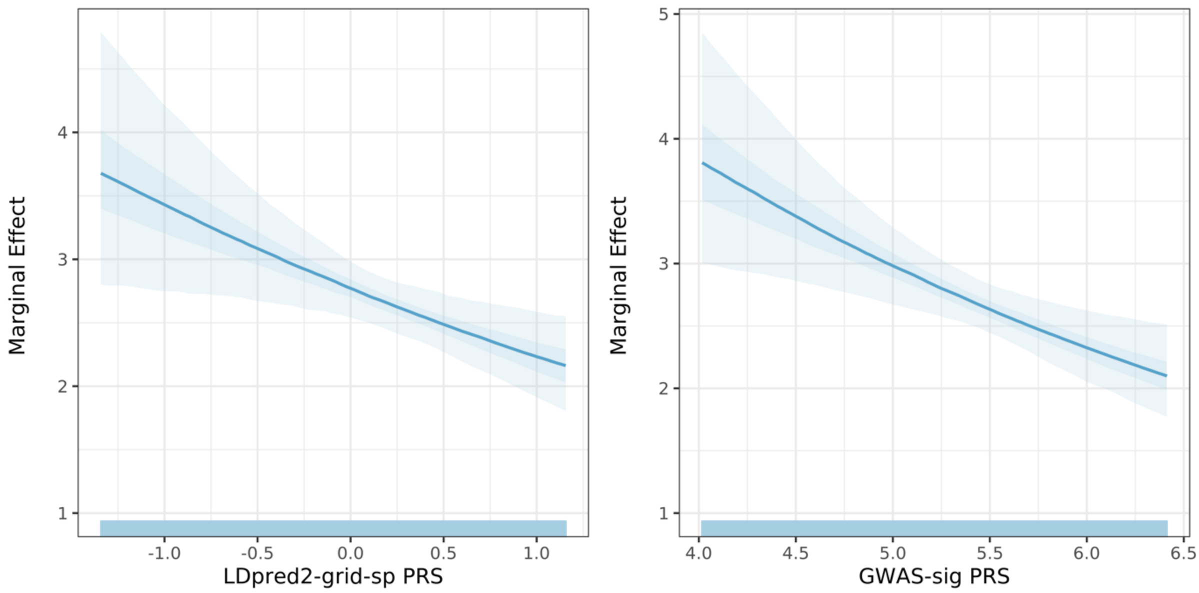

**Figure S18. Marginal effect of QCancer-10 risk score in interaction with PRS in male QCancer-10+LDP and QCancer-10+GWS models**

QCancer-10+LDP models for males after adjustment for optimism:

LP = 0.9783043*LDpred2-grid-sp + 0.976795*QCancer-10

Baseline survival function
5 years: 0.9954855
6 years: 0.9944618
7 years: 0.9934987
8 years: 0.9924138

QCancer-10+LDP models for females after adjustment for optimism:

LP = 0.2962966*$(\text{LDpred2-grid-sp}+1.4)^2$ + 0.7871845*(QCancer-10+0.8)

Baseline survival function
5 years: 0.9966845
6 years: 0.9959385
7 years: 0.9951428
8 years: 0.9944463

QCancer-10+GWS models for males after adjustment for optimism:

LP = 0.8083617*GWAS-sig + 0.9896891*QCancer-10

Baseline survival function
5 years: 0.9953175
6 years: 0.9942564
7 years: 0.9932547
8 years: 0.9921095

QCancer-10+GWS models for females after adjustment for optimism:

LP = 0.6610515*GWAS-sig + 0.7995745*QCancer-10

Baseline survival function
5 years: 0.9965650
6 years: 0.9957923
7 years: 0.9949641
8 years: 0.9942322

**Table S13. Sensitivity of QCancer-10+GWS models for CRC diagnosis over 5 years of follow-up across top 25 centiles of absolute risk in males and females**

| Centiles | Population per centile | Absolute 5-year risk centile cut-off (%) | Cases per centile | Cumulative % cases based on absolute risk (sensitivity) |
|---|---|---|---|---|
| **Men** | | | | |
| 1 | 1960 | 2.46 | 74 | 3.9 |
| 2 | 1961 | 2.15 | 65 | 7.3 |
| 3 | 1961 | 1.97 | 44 | 9.6 |
| 4 | 1961 | 1.83 | 54 | 12.4 |
| 5 | 1961 | 1.72 | 47 | 14.9 |
| 6 | 1961 | 1.63 | 39 | 17.0 |
| 7 | 1961 | 1.56 | 53 | 19.8 |
| 8 | 1961 | 1.50 | 37 | 21.8 |
| 9 | 1961 | 1.44 | 43 | 24.1 |
| 10 | 1961 | 1.39 | 47 | 26.6 |
| 11 | 1961 | 1.34 | 36 | 28.5 |
| 12 | 1960 | 1.30 | 42 | 30.7 |
| 13 | 1961 | 1.27 | 27 | 32.1 |
| 14 | 1961 | 1.23 | 47 | 34.6 |
| 15 | 1961 | 1.20 | 30 | 36.2 |
| 16 | 1961 | 1.17 | 28 | 37.7 |
| 17 | 1961 | 1.14 | 37 | 39.7 |
| 18 | 1961 | 1.11 | 22 | 40.9 |
| 19 | 1961 | 1.08 | 35 | 42.7 |
| 20 | 1961 | 1.06 | 39 | 44.8 |
| 21 | 1961 | 1.03 | 24 | 46.1 |
| 22 | 1961 | 1.01 | 31 | 47.7 |
| 23 | 1960 | 0.99 | 31 | 49.3 |
| 24 | 1961 | 0.96 | 32 | 51.0 |
| 25 | 1961 | 0.94 | 32 | 52.7 |
| **Women** | | | | |
| 1 | 2384 | 1.18 | 37 | 2.5 |
| 2 | 2385 | 1.06 | 39 | 5.2 |
| 3 | 2385 | 0.98 | 35 | 7.6 |
| 4 | 2385 | 0.93 | 31 | 9.7 |
| 5 | 2385 | 0.88 | 28 | 11.6 |
| 6 | 2385 | 0.85 | 30 | 13.7 |
| 7 | 2385 | 0.82 | 29 | 15.7 |
| 8 | 2385 | 0.79 | 34 | 18.0 |
| 9 | 2385 | 0.77 | 24 | 19.6 |
| 10 | 2385 | 0.75 | 25 | 21.3 |
| 11 | 2385 | 0.73 | 30 | 23.4 |
| 12 | 2385 | 0.71 | 35 | 25.8 |
| 13 | 2385 | 0.70 | 28 | 27.7 |
| 14 | 2385 | 0.68 | 20 | 29.1 |
| 15 | 2385 | 0.67 | 22 | 30.6 |
| 16 | 2385 | 0.65 | 21 | 32.0 |
| 17 | 2385 | 0.64 | 25 | 33.7 |
| 18 | 2385 | 0.63 | 22 | 35.2 |
| 19 | 2385 | 0.62 | 26 | 37.0 |
| 20 | 2385 | 0.61 | 23 | 38.6 |
| 21 | 2385 | 0.60 | 27 | 40.5 |
| 22 | 2385 | 0.59 | 26 | 42.3 |
| 23 | 2385 | 0.57 | 22 | 43.8 |
| 24 | 2385 | 0.57 | 20 | 45.2 |
| 25 | 2385 | 0.56 | 21 | 46.6 |

**Table S14. Sensitivity of QCancer-10 for CRC diagnosis over 5 years of follow-up across top 25 centiles of absolute risk in men and women.** Calculated following recalibration of the QCancer-10 model.

| Centiles | Population per centile | Absolute 5-year risk centile cut-off (%) | Cases per centile | Cumulative % cases based on absolute risk (sensitivity) |
|---|---|---|---|---|
| **Men** | | | | |
| 1 | 1960 | 1.90 | 49 | 2.6 |
| 2 | 1961 | 1.71 | 44 | 4.9 |
| 3 | 1961 | 1.60 | 51 | 7.6 |
| 4 | 1961 | 1.53 | 48 | 10.1 |
| 5 | 1961 | 1.47 | 51 | 12.8 |
| 6 | 1961 | 1.42 | 43 | 15.1 |
| 7 | 1961 | 1.38 | 35 | 16.9 |
| 8 | 1961 | 1.34 | 52 | 19.7 |
| 9 | 1961 | 1.31 | 36 | 21.6 |
| 10 | 1961 | 1.28 | 41 | 23.7 |
| 11 | 1961 | 1.25 | 37 | 25.7 |
| 12 | 1960 | 1.22 | 42 | 27.9 |
| 13 | 1961 | 1.20 | 38 | 29.9 |
| 14 | 1961 | 1.18 | 28 | 31.4 |
| 15 | 1961 | 1.16 | 39 | 33.5 |
| 16 | 1961 | 1.14 | 28 | 34.9 |
| 17 | 1961 | 1.12 | 36 | 36.8 |
| 18 | 1961 | 1.10 | 33 | 38.6 |
| 19 | 1961 | 1.08 | 27 | 40.0 |
| 20 | 1961 | 1.07 | 23 | 41.2 |
| 21 | 1961 | 1.05 | 27 | 42.6 |
| 22 | 1961 | 1.03 | 31 | 44.3 |
| 23 | 1960 | 1.01 | 25 | 45.6 |
| 24 | 1961 | 1.00 | 36 | 47.5 |
| 25 | 1961 | 0.98 | 22 | 48.7 |
| **Women** | | | | |
| 1 | 2336 | 1.10 | 24 | 1.6 |
| 2 | 2344 | 0.98 | 38 | 4.3 |
| 3 | 2364 | 0.91 | 22 | 5.8 |
| 4 | 2422 | 0.86 | 21 | 7.2 |
| 5 | 2375 | 0.82 | 34 | 9.5 |
| 6 | 2203 | 0.80 | 18 | 10.8 |
| 7 | 2598 | 0.78 | 24 | 12.4 |
| 8 | 1827 | 0.76 | 25 | 14.1 |
| 9 | 2991 | 0.75 | 24 | 15.8 |
| 10 | 900 | 0.74 | 8 | 16.3 |
| 11 | 3846 | 0.72 | 38 | 18.9 |
| 12 | 2392 | 0.71 | 22 | 20.4 |
| 13 | 1543 | 0.70 | 20 | 21.8 |
| 14 | 2417 | 0.69 | 35 | 24.2 |
| 15 | 3216 | 0.68 | 33 | 26.5 |
| 16 | 2294 | 0.67 | 26 | 28.3 |
| 17 | 2328 | 0.66 | 23 | 29.8 |
| 18 | 2499 | 0.65 | 16 | 30.9 |
| 19 | 2306 | 0.64 | 23 | 32.5 |
| 20 | 2434 | 0.63 | 30 | 34.6 |
| 21 | 2418 | 0.62 | 15 | 35.6 |
| 22 | 2058 | 0.61 | 27 | 37.4 |
| 23 | 2388 | 0.60 | 16 | 38.5 |
| 24 | 2542 | 0.59 | 19 | 39.8 |
| 25 | 2539 | 0.58 | 27 | 41.7 |

**Table S15. Sensitivity of QCancer-10+LDP across top 25 centiles of relative risk.** Risk is calculated relative to an individual of the same age and sex, of white-British ethnicity, with no CRC risk factors, BMI of 25, mean Townsend Deprivation Score, and mean PRS.

| Centiles | Population per centile | Age-sex relative risk centile cut-off (%) | Cases per centile | Cumulative % cases based on relative risk (sensitivity) |
|---|---|---|---|---|
| **Men** | | | | |
| 1 | 1960 | 4.81 | 55 | 2.9 |
| 2 | 1961 | 4.14 | 49 | 5.5 |
| 3 | 1961 | 3.78 | 36 | 7.4 |
| 4 | 1961 | 3.51 | 47 | 9.9 |
| 5 | 1961 | 3.32 | 32 | 11.6 |
| 6 | 1961 | 3.15 | 35 | 13.4 |
| 7 | 1961 | 3.01 | 38 | 15.4 |
| 8 | 1961 | 2.89 | 29 | 16.9 |
| 9 | 1961 | 2.79 | 40 | 19.0 |
| 10 | 1961 | 2.70 | 31 | 20.6 |
| 11 | 1961 | 2.62 | 36 | 22.5 |
| 12 | 1960 | 2.55 | 35 | 24.3 |
| 13 | 1961 | 2.48 | 40 | 26.4 |
| 14 | 1961 | 2.42 | 27 | 27.8 |
| 15 | 1961 | 2.36 | 27 | 29.2 |
| 16 | 1961 | 2.31 | 27 | 30.6 |
| 17 | 1961 | 2.26 | 22 | 31.8 |
| 18 | 1961 | 2.21 | 31 | 33.4 |
| 19 | 1961 | 2.17 | 26 | 34.8 |
| 20 | 1961 | 2.12 | 29 | 36.3 |
| 21 | 1961 | 2.08 | 28 | 37.8 |
| 22 | 1961 | 2.04 | 33 | 39.5 |
| 23 | 1960 | 2.01 | 30 | 41.1 |
| 24 | 1961 | 1.97 | 29 | 42.6 |
| 25 | 1961 | 1.94 | 29 | 44.1 |
| **Women** | | | | |
| 1 | 2199 | 3.89 | 46 | 3.2 |
| 2 | 2570 | 3.26 | 34 | 5.5 |
| 3 | 2384 | 2.90 | 41 | 8.3 |
| 4 | 2385 | 2.66 | 28 | 10.2 |
| 5 | 2386 | 2.48 | 29 | 12.2 |
| 6 | 2385 | 2.34 | 29 | 14.2 |
| 7 | 2385 | 2.23 | 25 | 15.9 |
| 8 | 2384 | 2.14 | 22 | 17.4 |
| 9 | 2386 | 2.06 | 26 | 19.2 |
| 10 | 2385 | 1.99 | 27 | 21.1 |
| 11 | 2385 | 1.92 | 26 | 22.9 |
| 12 | 2384 | 1.86 | 33 | 25.2 |
| 13 | 2385 | 1.81 | 21 | 26.6 |
| 14 | 2385 | 1.77 | 17 | 27.8 |
| 15 | 2386 | 1.72 | 14 | 28.8 |
| 16 | 2385 | 1.69 | 15 | 29.8 |
| 17 | 2385 | 1.65 | 17 | 31.0 |
| 18 | 2384 | 1.62 | 20 | 32.4 |
| 19 | 2386 | 1.58 | 19 | 33.7 |
| 20 | 2384 | 1.55 | 25 | 35.4 |
| 21 | 2385 | 1.52 | 18 | 36.6 |
| 22 | 2386 | 1.50 | 20 | 38.0 |
| 23 | 2385 | 1.47 | 21 | 39.4 |
| 24 | 2385 | 1.45 | 21 | 40.8 |
| 25 | 2385 | 1.42 | 19 | 42.1 |

**Table S16. Sensitivity of QCancer-10+GWS across top 25 centiles of relative risk.** Risk is calculated relative to an individual of the same age and sex, of white-British ethnicity, with no CRC risk factors, BMI of 25, mean Townsend Deprivation Score, and mean PRS.

| Centiles | Population per centile | Age-sex relative risk centile cut-off (%) | Cases per centile | Cumulative % cases based on relative risk (sensitivity) |
|---|---|---|---|---|
| **Men** | | | | |
| 1 | 1960 | 4.08 | 36 | 1.9 |
| 2 | 1961 | 3.54 | 45 | 4.3 |
| 3 | 1961 | 3.25 | 47 | 6.8 |
| 4 | 1961 | 3.06 | 41 | 9.0 |
| 5 | 1961 | 2.90 | 33 | 10.7 |
| 6 | 1961 | 2.78 | 33 | 12.4 |
| 7 | 1961 | 2.68 | 33 | 14.1 |
| 8 | 1961 | 2.58 | 36 | 16.0 |
| 9 | 1961 | 2.50 | 38 | 18.0 |
| 10 | 1961 | 2.43 | 43 | 20.3 |
| 11 | 1961 | 2.37 | 30 | 21.9 |
| 12 | 1960 | 2.31 | 26 | 23.3 |
| 13 | 1961 | 2.26 | 23 | 24.5 |
| 14 | 1961 | 2.21 | 26 | 25.9 |
| 15 | 1961 | 2.16 | 23 | 27.1 |
| 16 | 1961 | 2.12 | 18 | 28.0 |
| 17 | 1961 | 2.08 | 36 | 29.9 |
| 18 | 1961 | 2.04 | 25 | 31.2 |
| 19 | 1961 | 2.01 | 25 | 32.5 |
| 20 | 1961 | 1.98 | 28 | 34.0 |
| 21 | 1961 | 1.94 | 25 | 35.3 |
| 22 | 1961 | 1.91 | 25 | 36.6 |
| 23 | 1960 | 1.88 | 26 | 38.0 |
| 24 | 1961 | 1.86 | 24 | 39.3 |
| 25 | 1961 | 1.83 | 21 | 40.4 |
| **Women** | | | | |
| 1 | 2383 | 2.63 | 19 | 1.3 |
| 2 | 2386 | 2.36 | 35 | 3.7 |
| 3 | 2384 | 2.23 | 22 | 5.2 |
| 4 | 2386 | 2.12 | 24 | 6.8 |
| 5 | 2385 | 2.02 | 20 | 8.2 |
| 6 | 2385 | 1.95 | 22 | 9.7 |
| 7 | 2384 | 1.89 | 19 | 11.0 |
| 8 | 2385 | 1.84 | 20 | 12.4 |
| 9 | 2385 | 1.79 | 17 | 13.6 |
| 10 | 2385 | 1.75 | 33 | 15.9 |
| 11 | 2386 | 1.72 | 26 | 17.7 |
| 12 | 2385 | 1.68 | 20 | 19.1 |
| 13 | 2385 | 1.65 | 18 | 20.3 |
| 14 | 2385 | 1.62 | 21 | 21.7 |
| 15 | 2385 | 1.60 | 25 | 23.4 |
| 16 | 2385 | 1.57 | 19 | 24.7 |
| 17 | 2385 | 1.55 | 22 | 26.2 |
| 18 | 2385 | 1.53 | 19 | 27.5 |
| 19 | 2385 | 1.51 | 32 | 29.7 |
| 20 | 2384 | 1.49 | 21 | 31.1 |
| 21 | 2386 | 1.47 | 21 | 32.5 |
| 22 | 2384 | 1.45 | 21 | 33.9 |
| 23 | 2386 | 1.43 | 17 | 35.1 |
| 24 | 2384 | 1.41 | 19 | 36.4 |
| 25 | 2386 | 1.40 | 13 | 37.3 |

**Table S17. Fold-increase in absolute risk between 95th centile and median risk for QCancer-10+LDP, QCancer-10+GWS and QCancer-10 models**

| | QCancer-10+LDP | QCancer-10+GWS | QCancer-10 |
|---|---|---|---|
| Males | 3.49 | 3.14 | 2.37 |
| Females | 2.75 | 2.37 | 2.06 |

**Table S18. Percentage of population and cases with relative risk > 2.2 for QCancer-10+LDP, QCancer-10+GWS and QCancer-10 models**

| | QCancer-10+LDP | | QCancer-10+GWS | | QCancer-10 | |
|---|---|---|---|---|---|---|
| | Males | Females | Males | Females | Males | Females |
| % population with RR > 2.2 | 18.2 | 7.2 | 14.2 | 3.2 | 4.1 | 1.2 |
| % of individuals with RR > 2.2 without FDRCRC | 75.9 | 69.6 | 70.8 | 44.8 | 29.4 | 30.3 |
| % cases with RR > 2.2 | 34.0 | 16.5 | 26.3 | 6.0 | 4.9 | 1.6 |

**References**

1. Law PJ, Timofeeva M, Fernandez-Rozadilla C, et al. Association analyses identify 31 new risk loci for colorectal cancer susceptibility. *Nat Commun*. 2019;**10**(1):2154.DOI: 10.1038/s41467-019-09775-w
2. Aragon TJ. epitools: Epidemiology Tools. 2020 https://CRAN.R-project.org/package=epitools.
3. Therneau T. A Package for Survival Analysis in R. 2015 https://CRAN.R-project.org/package=survival.
4. Prive F, Arbel J, Vilhjalmsson BJ. LDpred2: better, faster, stronger. *Bioinformatics*. 2020;**36**(22-23):5424–31.DOI: 10.1093/bioinformatics/btaa1029
5. Collins GS, Ogundimu EO, Altman DG. Sample size considerations for the external validation of a multivariable prognostic model: a resampling study. *Stat Med*. 2016;**35**(2):214–26.DOI: 10.1002/sim.6787
6. Huyghe JR, Bien SA, Harrison TA, et al. Discovery of common and rare genetic risk variants for colorectal cancer. *Nat Genet*. 2019;**51**(1):76–87.DOI: 10.1038/s41588-018-0286-6
7. Bigdeli TB, Lee D, Webb BT, et al. A simple yet accurate correction for winner's curse can predict signals discovered in much larger genome scans. *Bioinformatics*. 2016;**32**(17):2598–603.DOI: 10.1093/bioinformatics/btw303
8. Prive F, Vilhjalmsson BJ, Aschard H, Blum MGB. Making the Most of Clumping and Thresholding for Polygenic Scores. *Am J Hum Genet*. 2019;**105**(6):1213–21.DOI: 10.1016/j.ajhg.2019.11.001
9. Lloyd-Jones LR, Zeng J, Sidorenko J, et al. Improved polygenic prediction by Bayesian multiple regression on summary statistics. *Nat Commun*. 2019;**10**.DOI: ARTN 5086
10.1038/s41467-019-12653-0
10. Choi SW, Mak TS, O'Reilly PF. Tutorial: a guide to performing polygenic risk score analyses. *Nat Protoc*. 2020;**15**(9):2759-72.DOI: 10.1038/s41596-020-0353-1
11. ClinRisk Ltd. QCancer®(15yr,colorectal). 2015 https://qcancer.org/15yr/colorectal/.
12. Usher-Smith JA, Harshfield A, Saunders CL, et al. External validation of risk prediction models for incident colorectal cancer using UK Biobank. *Br J Cancer*. 2018;**118**(5):750–9.DOI: 10.1038/bjc.2017.463
13. Riley RD, Snell KI, Ensor J, et al. Minimum sample size for developing a multivariable prediction model: PART II - binary and time-to-event outcomes. *Stat Med*. 2019;**38**(7):1276–96.DOI: 10.1002/sim.7992
14. Riley RD, Ensor J, Snell KIE, et al. Calculating the sample size required for developing a clinical prediction model. *BMJ*. 2020;**368**.DOI: ARTN m441
10.1136/bmj.m441
15. Royston P, Sauerbrei W. Improving the robustness of fractional polynomial models by preliminary covariate transformation: A pragmatic approach. *Comput Stat Data An*. 2007;**51**(9):4240–53.DOI: 10.1016/j.csda.2006.05.006
16. Prive F, Aschard H, Ziyatdinov A, Blum MGB. Efficient analysis of large-scale genome-wide data with two R packages: bigstatsr and bigsnpr. *Bioinformatics*. 2018;**34**(16):2781–7.DOI: 10.1093/bioinformatics/bty185
17. Harrell FE. rms: Regression Modeling Strategies. 2019 https://CRAN.R-project.org/package=rms.
18. Ambler G, Benner A. mfp: Multivariable Fractional Polynomials. 2015 https://CRAN.R-project.org/package=mfp.
19. Wickham H, Averick M, Bryan J, et al. Welcome to the tidyverse. *Journal of Open Source Software*. 2019;**4**(43):1686.DOI: https://doi.org/10.21105/joss.01686
20. Office for National Statistics. Cancer registration statistics, England [Available from: https://www.ons.gov.uk/peoplepopulationandcommunity/healthandsocialcare/conditionsanddiseases/datasets/cancerregistrationstatisticscancerregistrationstatisticsengland. Accessed September 2020].
